# Impact of Mandatory Grain Fortification with Folic Acid on Population Folate Levels and the Risk of Folate Deficiency and Insufficiency: A Systematic Review and Meta-Analysis

**DOI:** 10.1101/2025.08.22.25334177

**Authors:** Jessica Parker, Amanuel Kisho, Shuaikang Hou, Michael Goodman, Lauryn Cravens, Mary Ellen Grap, Helena Pachon

**Affiliations:** Department of Epidemiology, Rollins School of Public Health, Emory University, Atlanta, GA, USA; Food Fortification Initiative, Atlanta, GA, USA; College of Arts and Sciences, Georgia State University, Atlanta, GA, USA; Hubert Department of Global Health, Rollins School of Public Health, Emory University, Atlanta, USA

## Abstract

**Background:** Wheat flour, maize flour and rice (i.e. grains) fortification with folic acid is an important folate dietary source globally. There are no systematic reviews or meta-analyses evaluating the effect of mandatory grain fortification on folate insufficiency (using serum/plasma folate and red blood cell (RBC) folate), RBC folate levels or folate deficiency using RBC folate. This study assessed the effectiveness of mandatory grain fortification with folic acid on serum/plasma folate and red blood cell (RBC) folate levels and the risk of folate deficiency and insufficiency based on these biomarkers.

**Methods:** We searched PubMed and Embase with assistance from a professional library informationist. We selected studies from countries with mandatory grain fortification that include folic acid in standards if they reported primary pre- and post-fortification data on folate status outcomes. We ran meta-analyses in R using random effects models with results expressed as meta-differences of means (MDM) or meta-prevalence ratios (mPR) for continuous and binary outcomes respectively. All meta-estimates were accompanied by 95% confidence intervals (CI).

**Results:** We screened 4,311 documents, identifying 31 articles which reported folate status outcomes (22 reported mean serum/plasma or RBC folate, 18 reported prevalence of folate deficiency or insufficiency). About 19% of studies were conducted in low- or middle-income countries. Mandatory fortification improved folate status, albeit with considerable heterogeneity across studies (I^2^≥73%). For serum/plasma folate levels, the MDM across all studies was 15.0 nmol/L (95% CI: 9.4-20.5). For serum/plasma folate insufficiency and deficiency, the mPR (95% CI) estimates were 0.17 (0.08-0.37) and 0.08 (0.03-0.23), respectively. For red blood cell folate levels, the MDM was 329.4 nmol/L (95% CI 243.9-414.9). For RBC folate insufficiency and deficiency, mPRs (95% CIs) were 0.16 (0.08-0.30) and 0.05 (0.01-0.46), respectively.

**Conclusions:** Mandatory grain fortification with folic acid increases blood folate levels and decreases the risk of folate insufficiency and folate deficiency.

## Introduction

Folate (vitamin B9) is a nutrient responsible for vital physiological processes, including DNA synthesis and repair, red blood cell production, and metabolism.^1^ During pregnancy, folate facilitates neural tube formation, a stage of embryonic development crucial for brain and spine health.^2^ Neural tube defects occur when the neural tube fails to develop properly. The estimated global prevalence of neural tube defects is 20 cases per 10,000 births, with the highest rates disproportionately burdening low- or middle-income countries (LMICs).^3^

The World Health Organization defines folate deficiency as serum/plasma folate <6.8 nanomoles per liter and red blood cell folate <226.5 nanomoles per liter.^4^ The prevalence of folate deficiency among women of reproductive age exceeds 20% in LMICs and is less than 5% in high-income countries (HICs).^5^ Folate deficiency is associated with megaloblastic anemia and adverse pregnancy-related outcomes, including preterm delivery, spontaneous abortion, stillbirth, and congenital defects of the heart, brain, and spine.^4^

Even when women are not considered folate deficient, their folate levels may still be low enough to elevate the risk of having a pregnancy affected by a birth defect.^4^ This condition, known as folate insufficiency, is defined as red blood cell (RBC) folate values <906 nanomoles per liter^4^; there is currently no established serum/plasma folate sufficiency threshold for neural tube defect prevention.^6^ Given folate’s essential role in fetal cell development and function, particularly during early pregnancy, folate insufficiency poses a paramount public health concern, namely, an increased risk in having a pregnancy affected by a neural tube defect.^7^ In fact, maternal folate insufficiency is the most common risk factor for neural tube defects.^3^ The estimated prevalence of folate insufficiency is >40% in most countries regardless of income level.^5^

One of many interventions to prevent nutrient deficiencies and insufficiencies is mandatory food fortification at the country level, or the addition of vitamins and minerals to food.^8^ This improves the diet at a population level and reduces the prevalence of micronutrient malnutrition.^8^ Fortification with folic acid, a synthetic form of folate, has been associated with preventing congenital anomalies and birth defects, particularly neural tube defects (NTDs).^3,9–14^ One meta-analysis found an approximate 40% reduction in the odds of NTDs in low- or middle-income countries after fortification with folic acid.^14^

Ninety-three countries mandate wheat flour fortification alone or in combination with other grains (e.g., maize flour, rice)^15^; as of 2025, 70 of these countries include folate in mandatory grain fortification standards.^16^ Extensive evidence supports the safety of folic acid fortification, refuting concerns about potential adverse health effects such as masking of vitamin B12 deficiency and increasing cancer risk or mortality.^17–23^ Moreover, folic acid fortification has been shown to be a cost-effective intervention, as fortifying a country’s grain supply is less expensive than treating individuals with neural tube defects.^20,24–26^

Existing literature has broadened the understanding of how folic acid influences population folate levels by examining the impacts of voluntary fortification, supplementation, and efficacy trials.^14,27–29^ However, an unmet need remains to evaluate the real-world effectiveness of folic acid fortification of grains. Effectiveness of this intervention can be evaluated by focusing only on countries that have implemented mandatory grain fortification with folic acid. A 2010 literature review by Berry and colleagues identified an increase between pre- and post-fortification population serum/plasma and RBC folate levels in four countries – the USA, Canada, Chile, and Costa Rica.^30^

To our knowledge, no previous systematic review and meta-analysis examined the association between mandatory fortification of grains with folic acid and changes in folate concentrations, folate deficiency prevalence, and folate insufficiency prevalence. To close this knowledge gap we conducted a systematic review and meta-analysis of studies that reported pre- and post-fortification data in countries with mandatory grain fortification with folic acid to determine changes in serum/plasma folate concentration, RBC folate concentration, and the prevalence of folate deficiency and folate insufficiency based on each of the aforementioned biomarkers of folate status.

## Methods

The components and structure of this systematic review adhere to the PRISMA 2020 guidelines.^31^ The review was registered with Open Science Framework (OSF).^32^ As deidentified data were used throughout the review, no IRB approval was solicited.

### Data sources and search strategy

Systematic searches were conducted in PubMed and Embase on 4 October 2021 and 29 October 2021, respectively.^33,34^ The search used the adapted PICO protocol^35^:

Two searches were conducted in each database. The first search used the keywords: “folate or folic acid” AND “fortif* or enrich*.” The second search was created in consultation with a professional library informationist and used the following search string: (folate[tw] or “folic acid”[tw] OR “Folic Acid”[Mesh]) **AND** (“folic acid fortification”[tw] OR “food fortification*”[tw] OR fortify[tw] OR fortified[tw] OR fortification[tw] OR “food enrichment*”[tw] OR enrichment[tw] OR enriched[tw] OR supplement*[tw] OR concentration*[tw] OR status[tw] OR deficienc*[tw] OR “Food, Fortified”[Mesh] OR “Folic Acid Deficiency/prevention and control”[Mesh]) **AND** ( “wheat flour” [tw] OR “Flour”[Mesh] OR “corn flour”[tw] OR “maize flour”[tw] OR rice[tw] OR “Oryza”[Mesh]). All results from the four searches were downloaded as .RIS format files and uploaded into Covidence online system for further processing that included removal of duplicates, screening of titles and selection of full-text articles.^36^.

### Eligibility criteria

Studies from countries with mandatory fortification of wheat flour, maize flour and rice were eligible if the fortification standards included folic acid. Studies that reported data on voluntary fortification or supplementation were excluded. The review included studies that reported primary data. Studies had to report at least two data points for an outcome: one before the initiation of fortification in a country and one after fortification was implemented in the same country. Studies reporting any health outcome related to folic acid were included. Whereas the overall systematic review was designed to assess the relation of folic acid fortification with any health outcomes,^32^ the current analysis is focused specifically on serum/plasma folate and red blood cell (RBC) folate. Studies reporting health outcomes not exclusively related to folic acid were excluded, such as hemoglobin concentration, hematocrit percentage and prevalence of anemia.

Only effectiveness studies were considered eligible. For this reason, randomized trials, and animal or *in vitro* studies were excluded. Additionally, studies were excluded if the unit of analysis was food instead of folic acid content (e.g., grams per day of wheat flour or bread consumed), if biofortification was the primary intervention, if the only outcome reported was related to diet or consumption of folic acid, or if the study was a sensory or acceptability study. Mathematical modeling papers, letters to the editor, case reports, protocols, conference abstracts, and organizational statements were also excluded.

### Study selection

After removing duplicates, two researchers (MEG and HP) independently screened the titles and abstracts to identify candidates for full text review. The full texts of the studies were independently reviewed by two researchers (LC and HP) to determine if they met the inclusion criteria. For both screening and full text review, all disagreements were resolved through consensus.

### Data extraction and management

Relevant data were extracted from all studies that met the inclusion criteria. Two researchers (AK and JP) independently extracted the data, and any disagreements were resolved by a third researcher (HP) and/or through consensus. A data extraction form developed for the purposes of this study included three sections: overview, outcomes and risk of bias assessment.

The overview form recorded the last name of the first author and year of publication, the country where the study was conducted, whether the data were nationally representative, the year when mandatory fortification started, the year when post-fortification outcome data were collected, and the type of the grains (e.g., maize flour) fortified with folic acid. If available, the amount of the fortified food (e.g., bread, pasta) consumed at the beginning, and in the middle and end of the study, along with corresponding units (e.g., g/d) were recorded. The amount of folic acid added to the fortified grains, its units and any other nutrient added through fortification, were also documented.

The outcome form recorded the study population’s gender distribution and age (mean age or age range). If outcomes were reported for subgroups, the subgroup characteristics were captured in three ways: pregnancy or lactation status, health status (e.g., Crohn’s disease or cystic fibrosis), and other characteristics (e.g., age group, and race/ethnicity). For all outcomes, the following information was extracted: the outcome name, units (e.g., nanomoles per liter, or percent), and the pre- and post-fortification point estimates (e.g., mean, or median for continuous variables and proportion for binary variables) and measures of dispersion (e.g., standard deviation, standard error, or 95% confidence interval). Additionally, the sample size in the pre- and post-fortification period, a qualitative statement on whether the outcome improved, stayed the same, or worsened following fortification and the type of statistical test (if any) applied to examine the change in the parameter of interest. Whenever possible, the most adjusted outcome estimates were extracted.

### Risk of bias

The “Tool to Assess Risk of Bias in Cohort Studies” and the “Checklist for Analytical Cross Sectional Studies” were reviewed and modified to create the risk of bias section of the data extraction form.^37,38^ Different criteria were used to evaluate risk of bias, depending on study design.

Cohort studies (where samples from the same individuals were analyzed in the pre- and post-fortification periods) were assessed on the following six criteria: 1) exposed and unexposed groups in the study drawn from the same administrative database of patients presenting at same points of care over the same time frame; 2) study reported the amount of folic acid added to the food and the year of implementation; 3) study used validated instruments as measurement tools and data collectors were trained or educated in the use of the instruments with similar level of education, clinical or research experience; 4) study reported no missing outcome data or missing outcome data were balanced in numbers across fortification periods with similar reasons for missing data across groups; 5) study measured the intake of folate/folic acid from other sources (e.g., supplements) and also measured the intake of the food that was fortified before and after fortification; and 6) study used matching or stratification or multivariate regression analysis to adjust for confounding factors.

The corresponding criteria for serial cross-sectional studies that collected data from different individuals before and after fortification were as follows: 1) study stated clear and specific inclusion and exclusion criteria prior to recruitment of study participants (e.g., risk, stage of disease progression); 2) study provided clear description of study population demographics (e.g., at least age and sex), location, and time period; 3) study stated the amount of folic acid added to the food and the year of implementation; 4) study measured the intake of folic acid from other sources (e.g., supplements) and the intake of the food that was fortified before and after fortification; 5) study used validated instruments as measurement tools and data collectors were trained or educated in the use of the instrument(s) with similar level of education, clinical or research experience; and 6) study used appropriate statistical tests such as regression analysis or stratification.

The likelihood of publication bias was assessed using funnel plots and Egger’s test across all studies.

### Data synthesis and analysis

Statistical analyses were conducted using R software.^39^ The data elements used in the analysis included the pre- vs. post-fortification differences of means and the corresponding standard errors (SE). If a study reported absolute values before and after fortification, the differences of means and SE were estimated using OpenEpi online statistical calculator.^40^ If outcomes were measured on different scales (e.g., micrograms per liter vs. nanograms per milliliter in another study), they were converted to the same unit of nanomoles per liter (nmol/L) by multiplying the value by 2.266. If a study provided a median and minimum/maximum values or an interquartile range, the sample mean and SE were estimated using previously proposed methodology.^41^ Studies that reported a geometric mean instead of an arithmetic mean were excluded.

For binary outcomes, the data elements used in the analysis included the pre- vs. post-fortification numerator and the corresponding denominator. Studies that reported the prevalence of individuals with serum/plasma folate or RBC folate below a cutoff value were included in the analysis. There are recommended cutoffs to assess plasma folate deficiency (<6.8 nmol/L) and insufficiency (<22.5 nmol/L) as well as RBC folate deficiency (<226.5 nmol/L) and insufficiency (<906 nmol/L); however, prevalences calculated using any cutoff values reported by study authors were included ^4,6,42^

Studies (or outcomes within them) were excluded if the number of individuals with folate deficiency or insufficiency and the total sample size measured were not included in the studies or were not calculable with data from the studies; if folate deficiency or insufficiency was not reported; and if they reported repetitive data (e.g. data from overlapping years from the same country were reported in two or more studies; in these cases, the most recently published studies were included).

Research suggests that serum/plasma folate levels <22.5 nmol/L indicate folate insufficiency in populations with adequate vitamin B12 status ^6^. Therefore, studies reporting the prevalence of folate insufficiency were reviewed to determine if they reported on the vitamin B12 status of the population. The study population was considered to have adequate vitamin B12 status if the prevalence of vitamin B12 deficiency was 10% or less, or if the mean or median vitamin B12 was more than 200 pmol/L for any time point. Studies that reported inadequate vitamin B12 status were excluded.

All meta-analyses were conducted using inverse variance random effects models with results expressed as meta-differences of means (MDM) for continuous outcomes and prevalence ratios (mPR) for binary outcomes. All meta-estimates were accompanied by the corresponding 95% confidence intervals (CI), tests for heterogeneity and I-squared (I^2^) statistics. Forest plots and funnel plots were generated using R software^39^. By convention, I^2^ values of ≥75% were used as the cutoff for considerable levels of heterogeneity.^43,44^

## Results

### Search results

The initial literature search yielded 6,682 studies from electronic databases (**Figure 1**). After deduplication, 4,311 articles were identified for title/abstract screening. After screening, 188 studies were included for data extraction.

**Figure 1:**
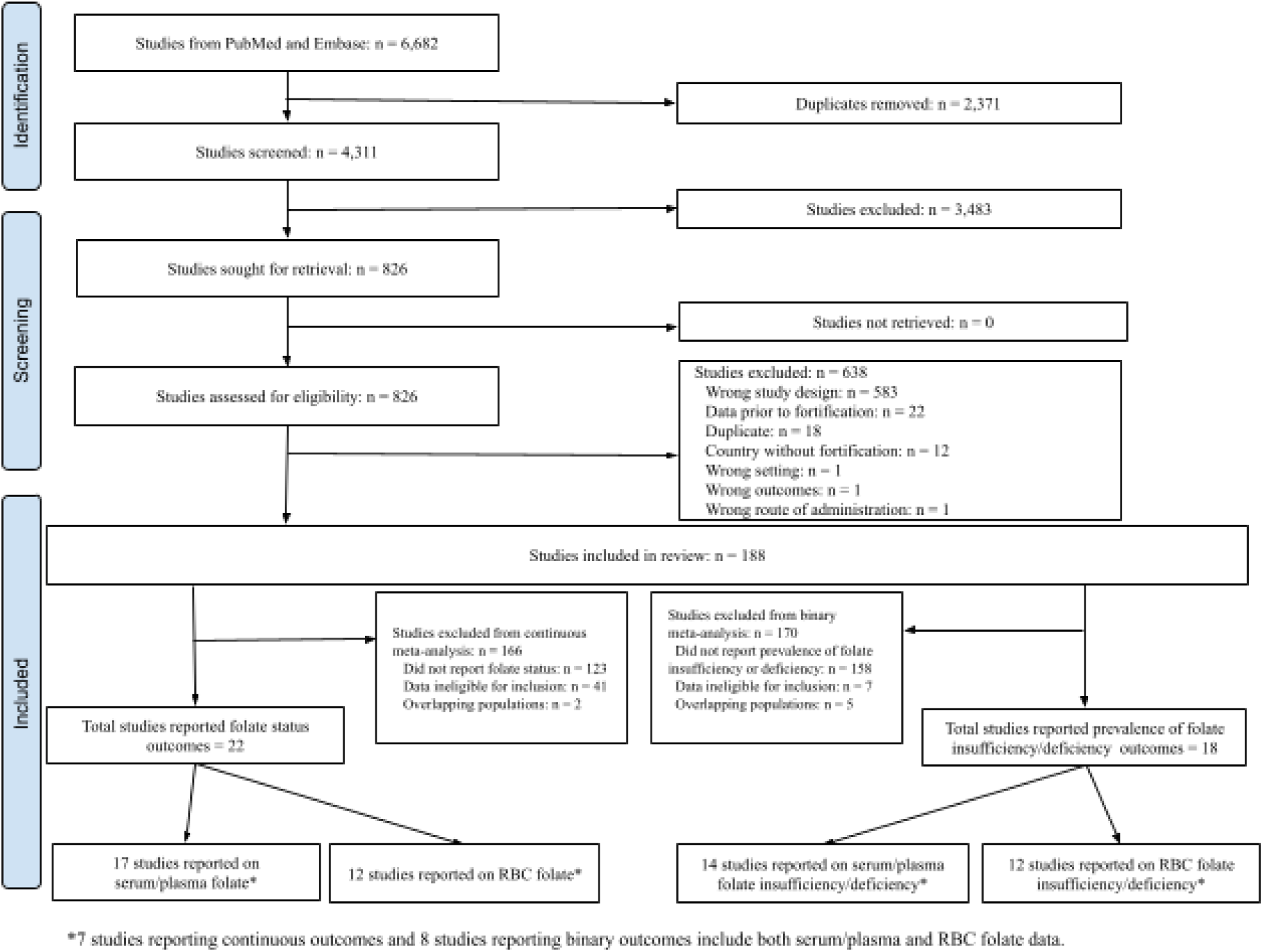
Summary of study selection: PRISMA Flow Chart for studies reporting on serum/plasma folate and red blood cell (RBC) folate continuous and binary outcomes and included in this analysis.^35^

### Study characteristics – continuous outcomes

Among 188 studies, 22 met the final inclusion criteria for continuous outcomes, namely reporting on folate status outcomes, using a unique data source, and complete outcome data (**Figure 1**), with 17 studies reporting results for serum/plasma folate and 12 studies reporting results for RBC folate. Seven studies presented data for both serum/plasma and RBC folate.

Taken together, the included studies provided data on 130,083 pre-fortification individuals and 90,414 post-fortification individuals with years of mandatory fortification initiation ranging from 1998 to 2011 (**Table 1**). With respect to geographic location, the data were obtained from 9 countries, including three LMIC (Iran, South Africa, Tanzania) and six HIC; the USA was the country with the most studies (n=8). There were 8 cohort studies and 14 were multiple-cross sectional studies. Out of 22 studies, 8 received a high-bias risk score, while 16 received a low-bias risk score (**Table 3**, **Table 4)**.

**Table 1:**
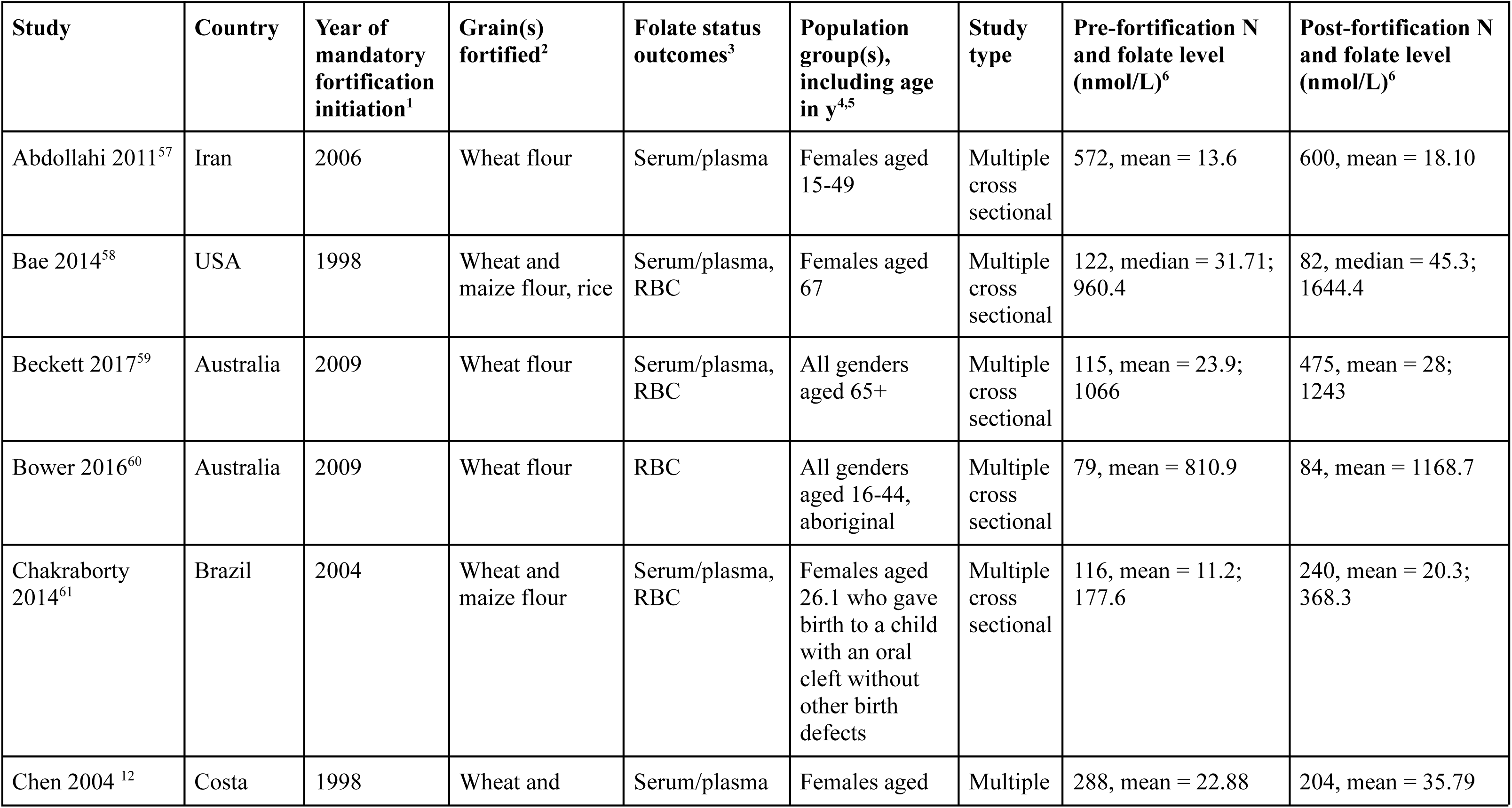
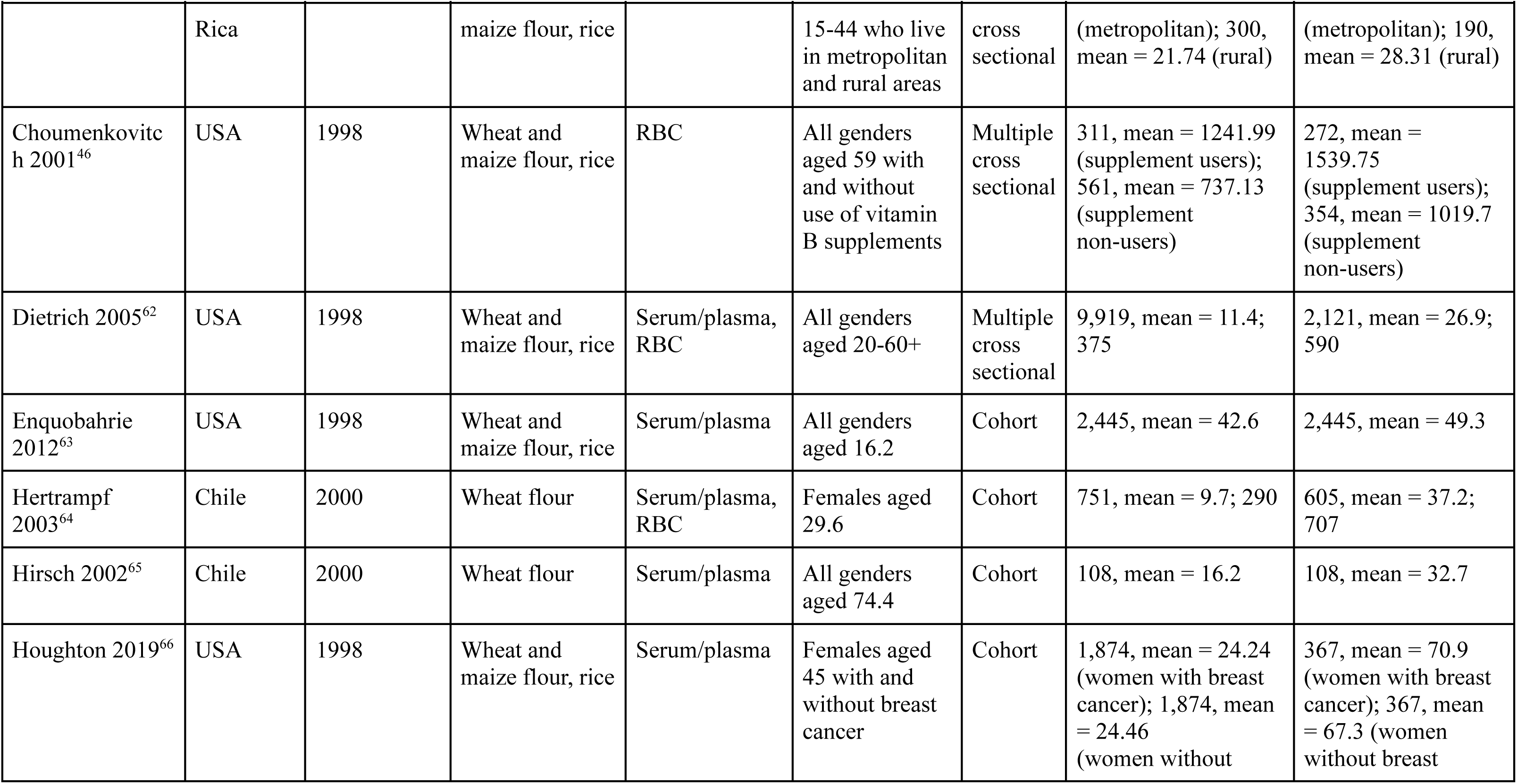
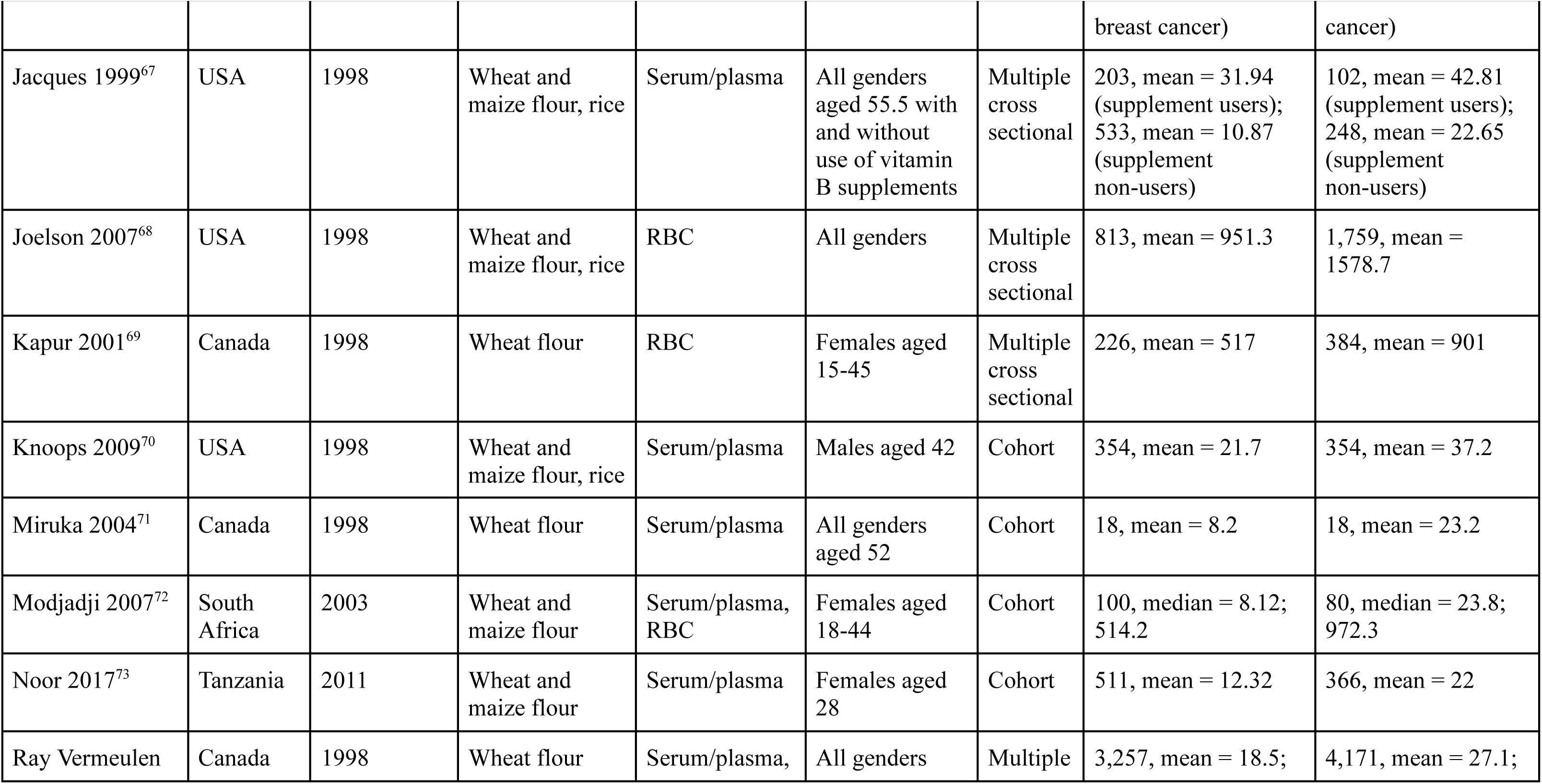
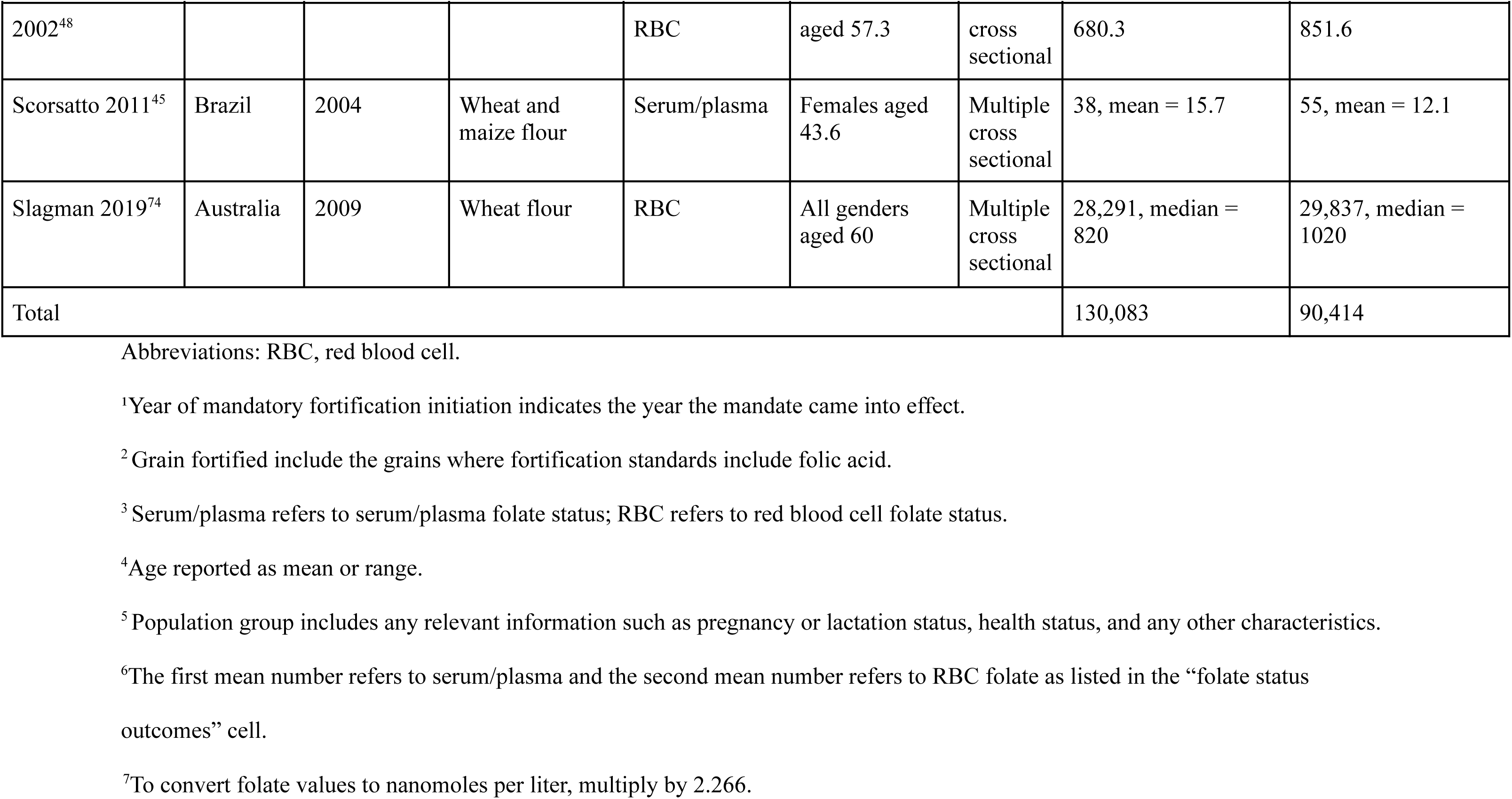
Summary of characteristics of each included study, review and meta-analysis of continuous folate status outcomes in countries with mandatory wheat flour, maize flour or rice fortification with folic acid (n=22).

| **Study** | **Country** | **Year of mandatory fortification initiation<sup>1</sup>** | **Grain(s) fortified<sup>2</sup>** | **Folate status outcomes<sup>3</sup>** | **Population group(s), including age in y<sup>4,5</sup>** | **Study type** | **Pre-fortification N and folate level (nmol/L)<sup>6</sup>** | **Post-fortification N and folate level (nmol/L)<sup>6</sup>** |
| --- | --- | --- | --- | --- | --- | --- | --- | --- |
| Abdollahi 2011<sup>57</sup> | Iran | 2006 | Wheat flour | Serum/plasma | Females aged 15-49 | Multiple cross sectional | 572, mean = 13.6 | 600, mean = 18.10 |
| Bae 2014<sup>58</sup> | USA | 1998 | Wheat and maize flour, rice | Serum/plasma, RBC | Females aged 67 | Multiple cross sectional | 122, median = 31.71; 960.4 | 82, median = 45.3; 1644.4 |
| Beckett 2017<sup>59</sup> | Australia | 2009 | Wheat flour | Serum/plasma, RBC | All genders aged 65+ | Multiple cross sectional | 115, mean = 23.9; 1066 | 475, mean = 28; 1243 |
| Bower 2016<sup>60</sup> | Australia | 2009 | Wheat flour | RBC | All genders aged 16-44, aboriginal | Multiple cross sectional | 79, mean = 810.9 | 84, mean = 1168.7 |
| Chakraborty 2014<sup>61</sup> | Brazil | 2004 | Wheat and maize flour | Serum/plasma, RBC | Females aged 26.1 who gave birth to a child with an oral cleft without other birth defects | Multiple cross sectional | 116, mean = 11.2; 177.6 | 240, mean = 20.3; 368.3 |
| Chen 2004<sup>12</sup> | Costa | 1998 | Wheat and | Serum/plasma | Females aged | Multiple | 288, mean = 22.88 | 204, mean = 35.79 |

| Study | Country | Year of mandatory fortification initiation<sup>1</sup> | Grain(s) fortified<sup>2</sup> | Folate status outcomes<sup>3</sup> | Population group(s), including age in y<sup>4,5</sup> | Study type | Pre-fortification N and folate level (nmol/L)<sup>6</sup> | Post-fortification N and folate level (nmol/L)<sup>6</sup> |
| --- | --- | --- | --- | --- | --- | --- | --- | --- |
|  | Rica |  | maize flour, rice |  | 15-44 who live in metropolitan and rural areas | cross sectional | (metropolitan); 300, mean = 21.74 (rural) | (metropolitan); 190, mean = 28.31 (rural) |
| Choumenkovitch 2001<sup>46</sup> | USA | 1998 | Wheat and maize flour, rice | RBC | All genders aged 59 with and without use of vitamin B supplements | Multiple cross sectional | 311, mean = 1241.99 (supplement users); 561, mean = 737.13 (supplement non-users) | 272, mean = 1539.75 (supplement users); 354, mean = 1019.7 (supplement non-users) |
| Dietrich 2005<sup>62</sup> | USA | 1998 | Wheat and maize flour, rice | Serum/plasma, RBC | All genders aged 20-60+ | Multiple cross sectional | 9,919, mean = 11.4; 375 | 2,121, mean = 26.9; 590 |
| Enquobahrie 2012<sup>63</sup> | USA | 1998 | Wheat and maize flour, rice | Serum/plasma | All genders aged 16.2 | Cohort | 2,445, mean = 42.6 | 2,445, mean = 49.3 |
| Hertrampf 2003<sup>64</sup> | Chile | 2000 | Wheat flour | Serum/plasma, RBC | Females aged 29.6 | Cohort | 751, mean = 9.7; 290 | 605, mean = 37.2; 707 |
| Hirsch 2002<sup>65</sup> | Chile | 2000 | Wheat flour | Serum/plasma | All genders aged 74.4 | Cohort | 108, mean = 16.2 | 108, mean = 32.7 |
| Houghton 2019<sup>66</sup> | USA | 1998 | Wheat and maize flour, rice | Serum/plasma | Females aged 45 with and without breast cancer | Cohort | 1,874, mean = 24.24 (women with breast cancer); 1,874, mean = 24.46 (women without breast cancer) | 367, mean = 70.9 (women with breast cancer); 367, mean = 67.3 (women without breast cancer) |
| Jacques 1999<sup>67</sup> | USA | 1998 | Wheat and maize flour, rice | Serum/plasma | All genders aged 55.5 with and without use of vitamin B supplements | Multiple cross sectional | 203, mean = 31.94 (supplement users); 533, mean = 10.87 (supplement non-users) | 102, mean = 42.81 (supplement users); 248, mean = 22.65 (supplement non-users) |
| Joelson 2007<sup>68</sup> | USA | 1998 | Wheat and maize flour, rice | RBC | All genders | Multiple cross sectional | 813, mean = 951.3 | 1,759, mean = 1578.7 |
| Kapur 2001<sup>69</sup> | Canada | 1998 | Wheat flour | RBC | Females aged 15-45 | Multiple cross sectional | 226, mean = 517 | 384, mean = 901 |
| Knoops 2009<sup>70</sup> | USA | 1998 | Wheat and maize flour, rice | Serum/plasma | Males aged 42 | Cohort | 354, mean = 21.7 | 354, mean = 37.2 |
| Miruka 2004<sup>71</sup> | Canada | 1998 | Wheat flour | Serum/plasma | All genders aged 52 | Cohort | 18, mean = 8.2 | 18, mean = 23.2 |
| Modjadji 2007<sup>72</sup> | South Africa | 2003 | Wheat and maize flour | Serum/plasma, RBC | Females aged 18-44 | Cohort | 100, median = 8.12; 514.2 | 80, median = 23.8; 972.3 |
| Noor 2017<sup>73</sup> | Tanzania | 2011 | Wheat and maize flour | Serum/plasma | Females aged 28 | Cohort | 511, mean = 12.32 | 366, mean = 22 |
| Ray Vermeulen | Canada | 1998 | Wheat flour | Serum/plasma, | All genders | Multiple | 3,257, mean = 18.5; | 4,171, mean = 27.1; |
| 2002<sup>48</sup> |  |  |  | RBC | aged 57.3 | cross sectional | 680.3 | 851.6 |
| Scorsatto 2011<sup>45</sup> | Brazil | 2004 | Wheat and maize flour | Serum/plasma | Females aged 43.6 | Multiple cross sectional | 38, mean = 15.7 | 55, mean = 12.1 |
| Slagman 2019<sup>74</sup> | Australia | 2009 | Wheat flour | RBC | All genders aged 60 | Multiple cross sectional | 28,291, median = 820 | 29,837, median = 1020 |
| Total |  |  |  |  |  |  | 130,083 | 90,414 |

Abbreviations: RBC, red blood cell.
<sup>1</sup>Year of mandatory fortification initiation indicates the year the mandate came into effect.
<sup>2</sup>Grain fortified include the grains where fortification standards include folic acid.
<sup>3</sup>Serum/plasma refers to serum/plasma folate status; RBC refers to red blood cell folate status.
<sup>4</sup>Age reported as mean or range.
<sup>5</sup>Population group includes any relevant information such as pregnancy or lactation status, health status, and any other characteristics.
<sup>6</sup>The first mean number refers to serum/plasma and the second mean number refers to RBC folate as listed in the “folate status outcomes” cell.
<sup>7</sup>To convert folate values to nanomoles per liter, multiply by 2.266.

### Study characteristics – binary outcomes

Of the 188 extracted studies, 18 studies met the final inclusion criteria (reporting on folate deficiency or insufficiency prevalence outcomes, using a unique data source, and complete outcome data) (**Figure 1**), with 14 studies reporting results for serum/plasma folate and 12 studies reporting results for RBC folate. Eight studies presented data for both RBC and serum/plasma folate.

Taken together, the included studies provided data on 50,511 pre-fortification individuals and 44,729 post-fortification individuals with years of mandatory fortification initiation ranging from 1997 to 2011 (**Table 2**). With respect to geographic location, the data were obtained from nine countries, including five LMIC (Brazil, Costa Rica, Iran, South Africa, Tanzania) and four HIC; the USA was the country with the most studies (n=13). There were nine cohort studies and 19 were multiple-cross sectional studies. Out of 18 studies, 10 received a high-bias risk score, while 8 received a low-bias risk score (**Table 3**, **Table 4**).

**Table 2:**
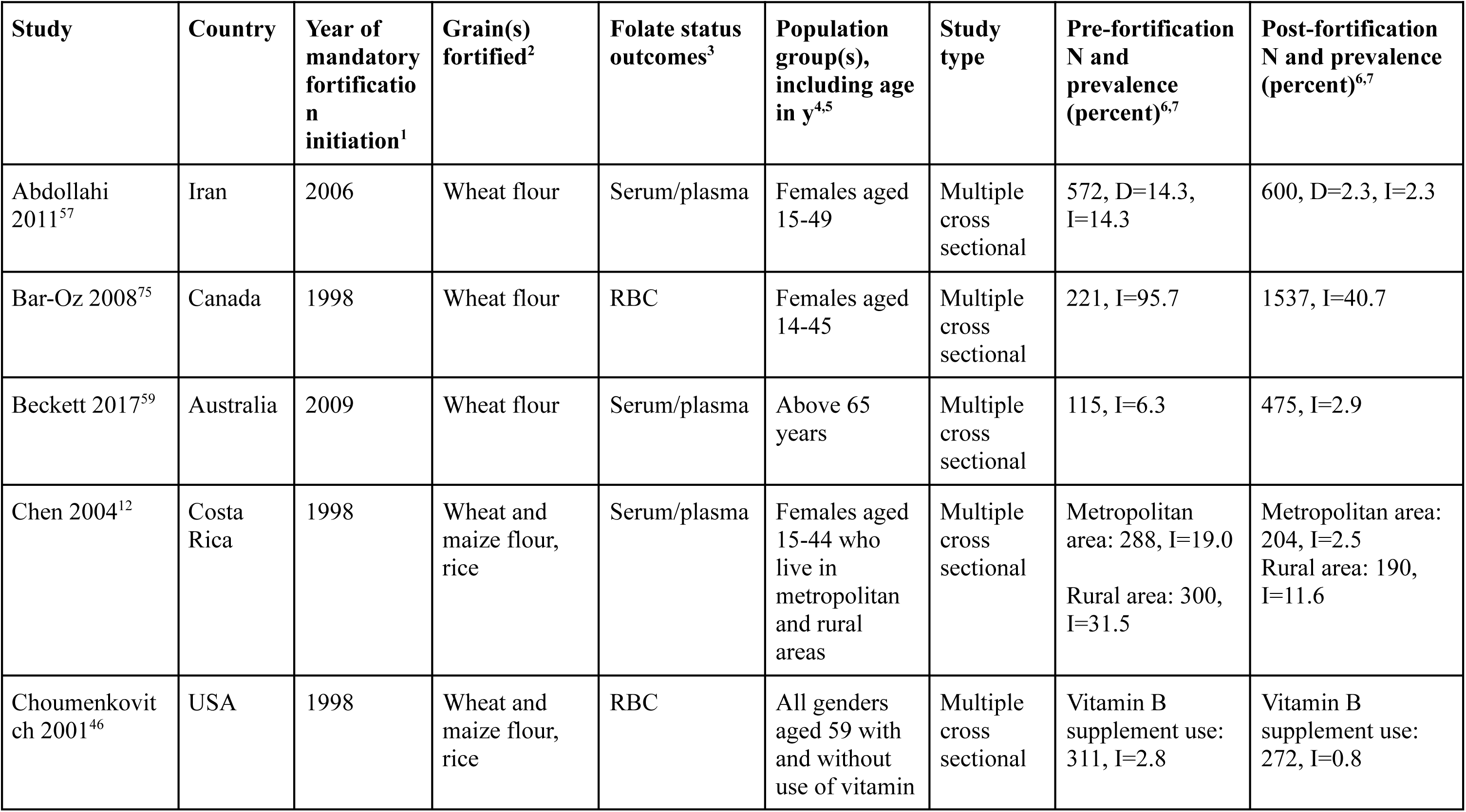
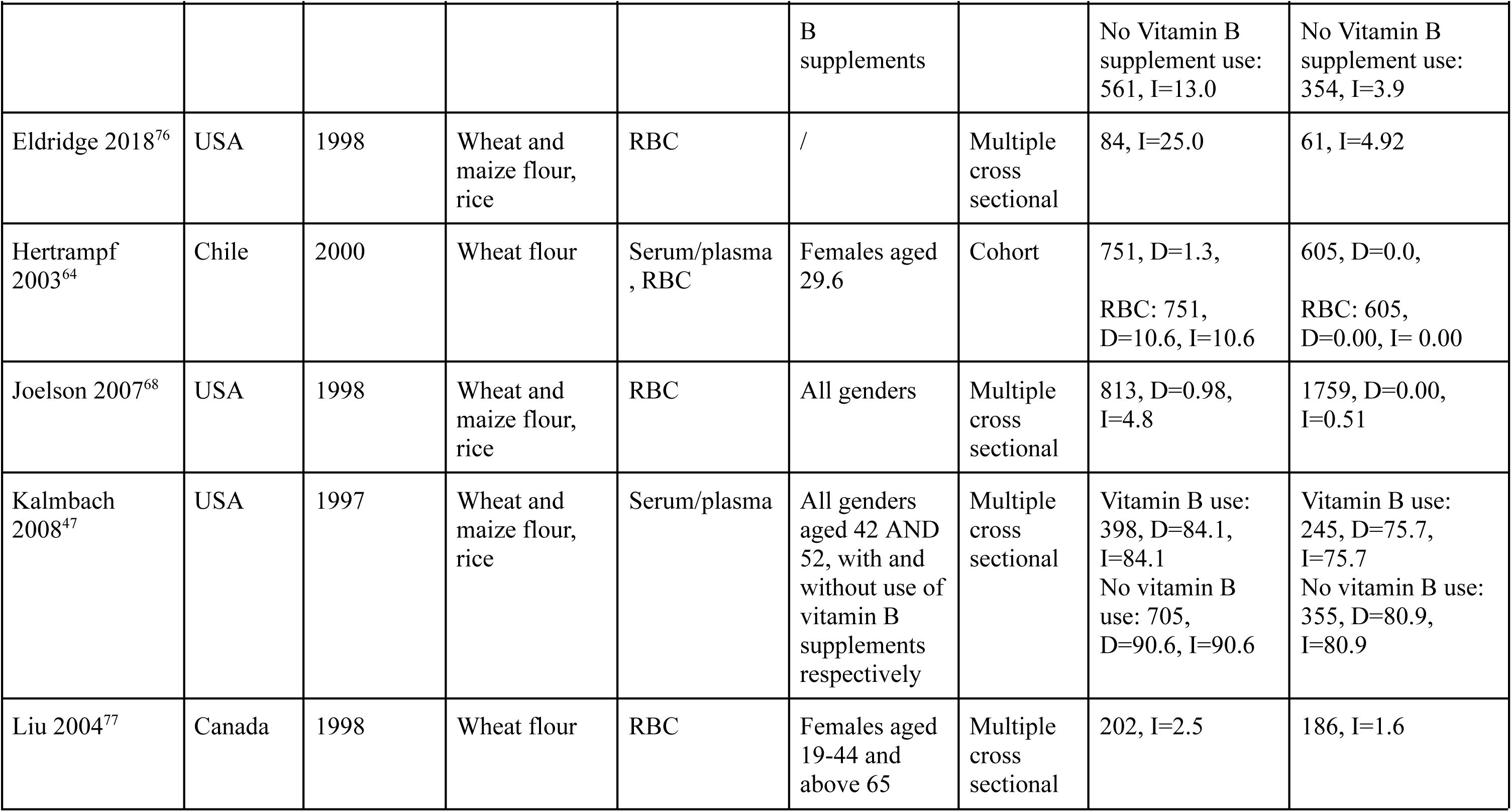
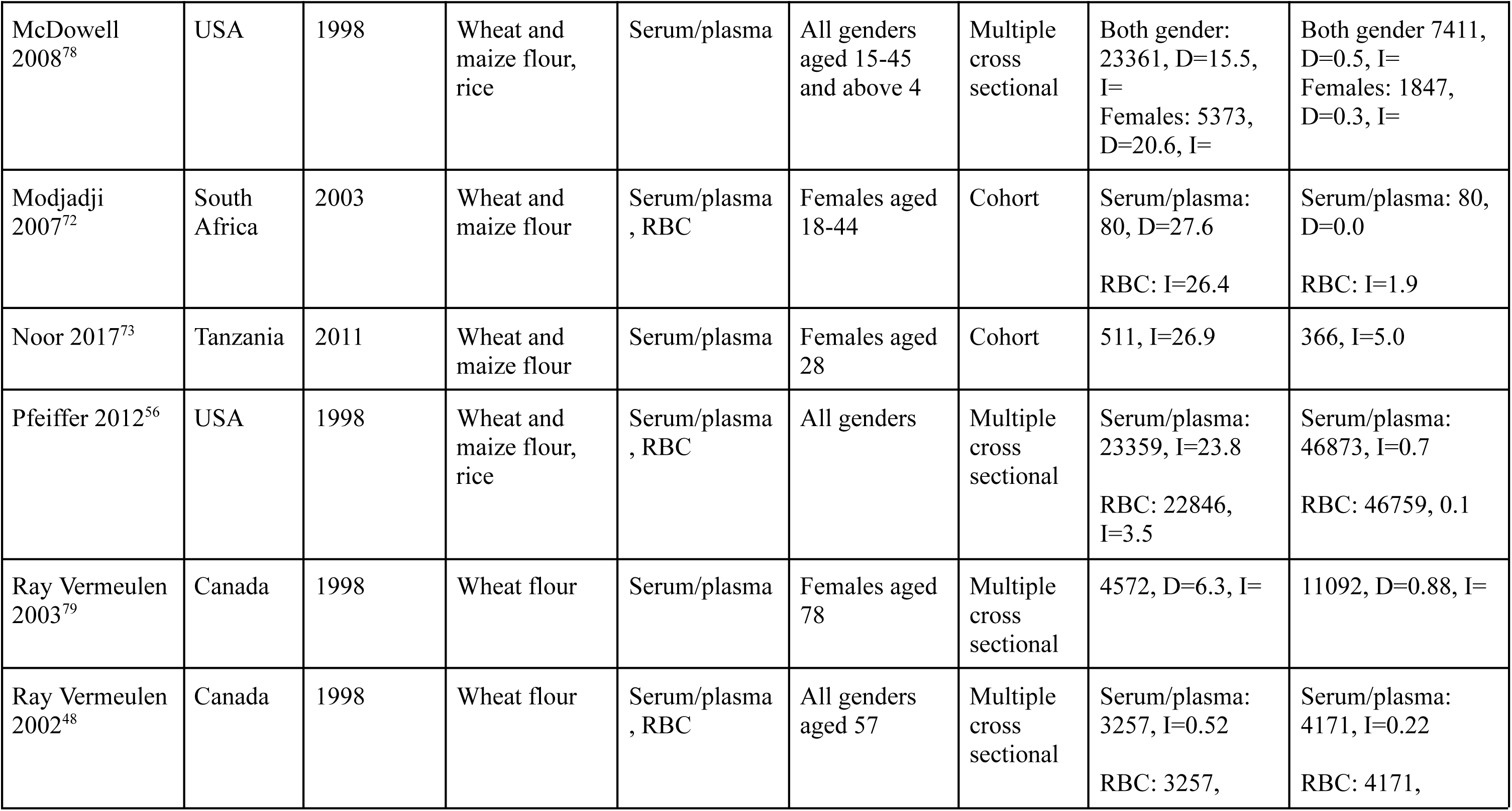
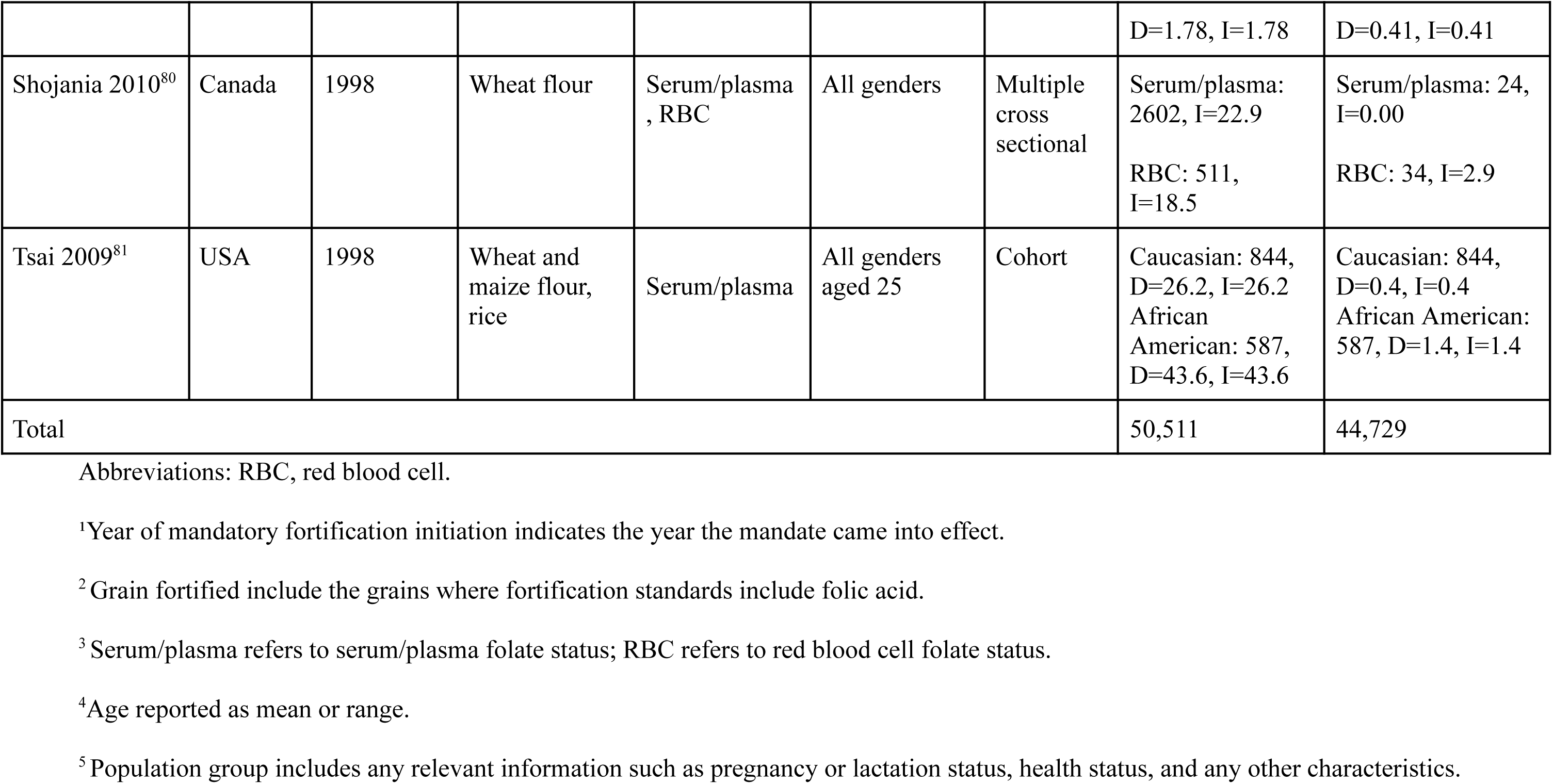
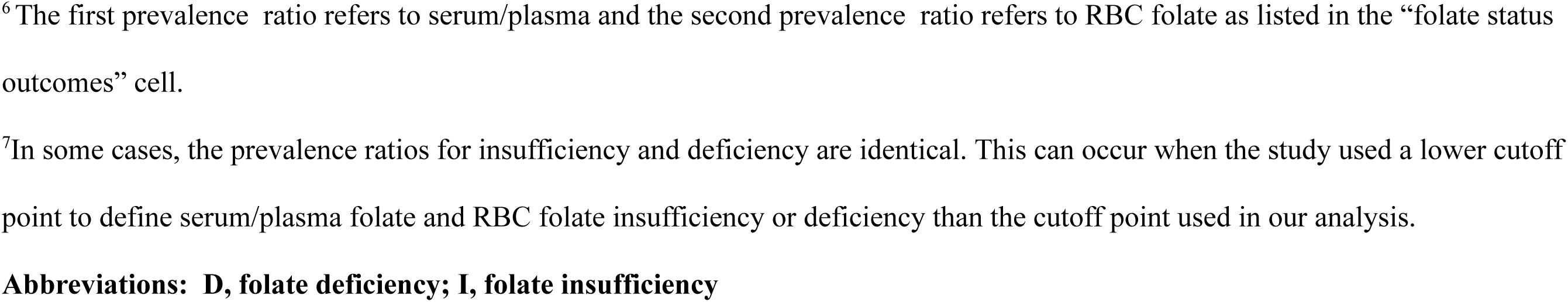
Summary of characteristics of each included study, review and meta-analysis of folate deficiency (d) or insufficiency (I) prevalence outcomes in countries with mandatory wheat flour, maize flour or rice fortification with folic acid (n=18).

**Table 3:** Risk of bias summary for multiple cross-sectional studies: review authors’ judgements about each risk of bias (ROB) item (n=23).

| Study | Continuous outcomes<sup>1</sup> | Binary outcomes<sup>2</sup> | Domain |  |  |  |  |  |  |
| --- | --- | --- | --- | --- | --- | --- | --- | --- | --- |
|  |  |  | Inclusion Criteria (1) | Subjects & Setting (2) | Exposure Measurement (3) | Confounders (4) | Outcome Measurement (5) | Statistical Analyses (6) | Overall ROB<sup>3</sup> |
| Abdollahi 2011 | Yes | Yes | Low | Low | Low | High | Low | Low | Low risk |
| Bar-Oz 2008 | No | Yes | Low | Low | Low | High | High | Low | Low risk |
| Bae 2014 | Yes | No | Low | High | Low | High | High | Low | High risk |
| Beckett 2017 | Yes | Yes | Low | Low | Low | Low | High | Low | Low risk |
| Bower 2016 | Yes | No | Low | Low | High | Low | High | Low | Low risk |
| Chakraborty 2014 | Yes | No | Low | Low | Low | High | High | Low | Low risk |
| Chen 2004 | Yes | Yes | Low | Low | Low | High | High | Low | Low risk |
| Choumenkovitch 2001 | Yes | Yes | Low | Low | Low | Low | High | Low | Low risk |
| Eldridge 2018 | No | Yes | Low | Low | High | High | High | Low | High risk |
|  |  |  | Inclusion Criteria (1) | Subjects & Setting (2) | Exposure Measurement (3) | Confounders (4) | Outcome Measurement (5) | Statistical Analyses (6) | Overall ROB<sup>3</sup> |
| Dietrich 2005 | Yes | No | Low | Low | High | Low | High | Low | Low risk |
| Jacques 1999 | Yes | No | Low | Low | Low | High | High | High | High risk |
| Joelson 2007 | Yes | Yes | Low | High | Low | High | High | Low | High risk |
| Kalmbach 2008 | No | Yes | Low | Low | Low | High | High | Low | Low risk |
| Kapur 2001 | Yes | No | Low | Low | Low | High | High | High | High risk |
| Liu 2004 | No | Yes | Low | Low | High | High | High | Low | High risk |
| McDowell 2008 | No | Yes | Low | Low | High | High | High | Low | High risk |
| Pfeiffer 2012 | No | Yes | Low | Low | High | High | High | Low | High risk |
| Ray Vermeulen 2002 | No | Yes | Low | Low | High | High | High | Low | High risk |
| Ray Vermeulen 2003 | No | Yes | Low | Low | High | High | High | Low | High risk |
| Scorsatto 2011 | Yes | No | Low | Low | Low | High | High | Low | Low risk |
|  |  |  | Inclusion Criteria (1) | Subjects & Setting (2) | Exposure Measurement (3) | Confounders (4) | Outcome Measurement (5) | Statistical Analyses (6) | Overall ROB<sup>3</sup> |
| Shojania 2010 | No | Yes | Low | High | Low | High | High | Low | High risk |
| Slagman 2019 | Yes | No | Low | Low | Low | High | High | Low | Low risk |

<sup>1</sup> This study reported continuous outcomes (e.g. mean red blood cell folate concentration).
<sup>2</sup> This study reported binary outcomes (e.g. prevalence of low red blood cell folate).
<sup>3</sup> Overall risk of bias was determined by dividing the number of criteria fulfilled by the total number of criteria (listed below) If 3 or more criteria were met, the ROB was considered low.
1. Were the criteria for inclusion in the sample clearly defined? 2. Were the study subjects and the setting described in detail? 3.
Was the exposure measured in a valid and reliable way? 4. Were confounding factors identified? 5. Was the outcome measured in a valid and reliable way? 6. Were the statistical analyses used documented?

**Table 4:** Risk of bias summary for cohort studies: review authors’ judgements about each risk of bias (ROB) item (n=9).

| Study | Continuous outcomes<sup>1</sup> | Binary outcomes<sup>2</sup> | Domain |  |  |  |  |  |  |
| --- | --- | --- | --- | --- | --- | --- | --- | --- | --- |
|  |  |  | Cohort Selection (1) | Exposure Measurement (2) | Outcome Measurement (3) | Cohort Follow-up (4) | Confounders (5) | Strategies for Confounding (6) | Overall ROB<sup>3</sup> |
| Enquobahrie 2012 | Yes | No | Low | Low | Low | Low | Low | Low | Low risk |
| Hertrampf 2003 | Yes | Yes | Low | Low | High | Low | Low | High | Low risk |
| Hirsch 2002 | Yes | No | Low | Low | High | Low | High | High | High risk |
| Houghton 2018 | Yes | No | Low | High | High | Low | Low | Low | Low risk |
| Knoops 2009 | Yes | No | Low | Low | High | Low | High | Low | Low risk |
| Miruka 2004 | Yes | No | Low | High | High | Low | High | Low | High risk |
| Modjadji 2007 | Yes | Yes | Low | High | High | Low | High | High | High risk |
|  |  |  | Cohort Selection (1) | Exposure Measurement (2) | Outcome Measurement (3) | Cohort Follow-up (4) | Confounders (5) | Strategies for Confounding (6) | Overall ROB<sup>3</sup> |
| Noor 2017 | Yes | Yes | Low | Low | Low | Low | Low | Low | Low risk |
| Tsai 2009 | No | Yes | Low | High | High | Low | High | Low | High risk |

<sup>1</sup> This study reported continuous outcomes (e.g. mean red blood cell folate concentration).
<sup>2</sup> This study reported binary outcomes (e.g. prevalence of low red blood cell folate).
<sup>3</sup> Overall risk of bias was determined by dividing the number of criteria fulfilled by the total number of criteria (listed below) If 3 or more criteria were met, the ROB was considered low.
1. Was selection of exposed and non-exposed cohorts drawn from the same population? 2. Was the exposure measured in a valid and reliable way? 3. Was the outcome measured in a valid and reliable way? 4. Was the follow up of the cohorts adequate? 5. Were confounding factors identified? 6. Were strategies to deal with confounding factors stated?

### Results of meta-analyses for serum/plasma folate: continuous and binary outcomes

Mandatory fortification of wheat flour, maize flour, or rice with folic acid led to an increase in serum/plasma folate levels in all but one study (**Figure 2**). The only exception was Scorsatto 2011^45^, which had a mean difference of -3.6 nmol/L. The MDM across all studies was 15.0 nmol/L (95% CI 9.4-20.5); however the results were very heterogeneous (I^2^=99%, p<0.001), indicating that the impact of fortification differed significantly across populations or settings. When stratified by risk of bias (**Supplementary Figure 1**), studies with a high risk of bias had a MDM of 13.1 nmol/L (95% CI 10.2-15.9), whereas studies with a low risk of bias had a MDM of 15.2 nmol/L (95% CI 7.1-23.4).

**Figure 2:**
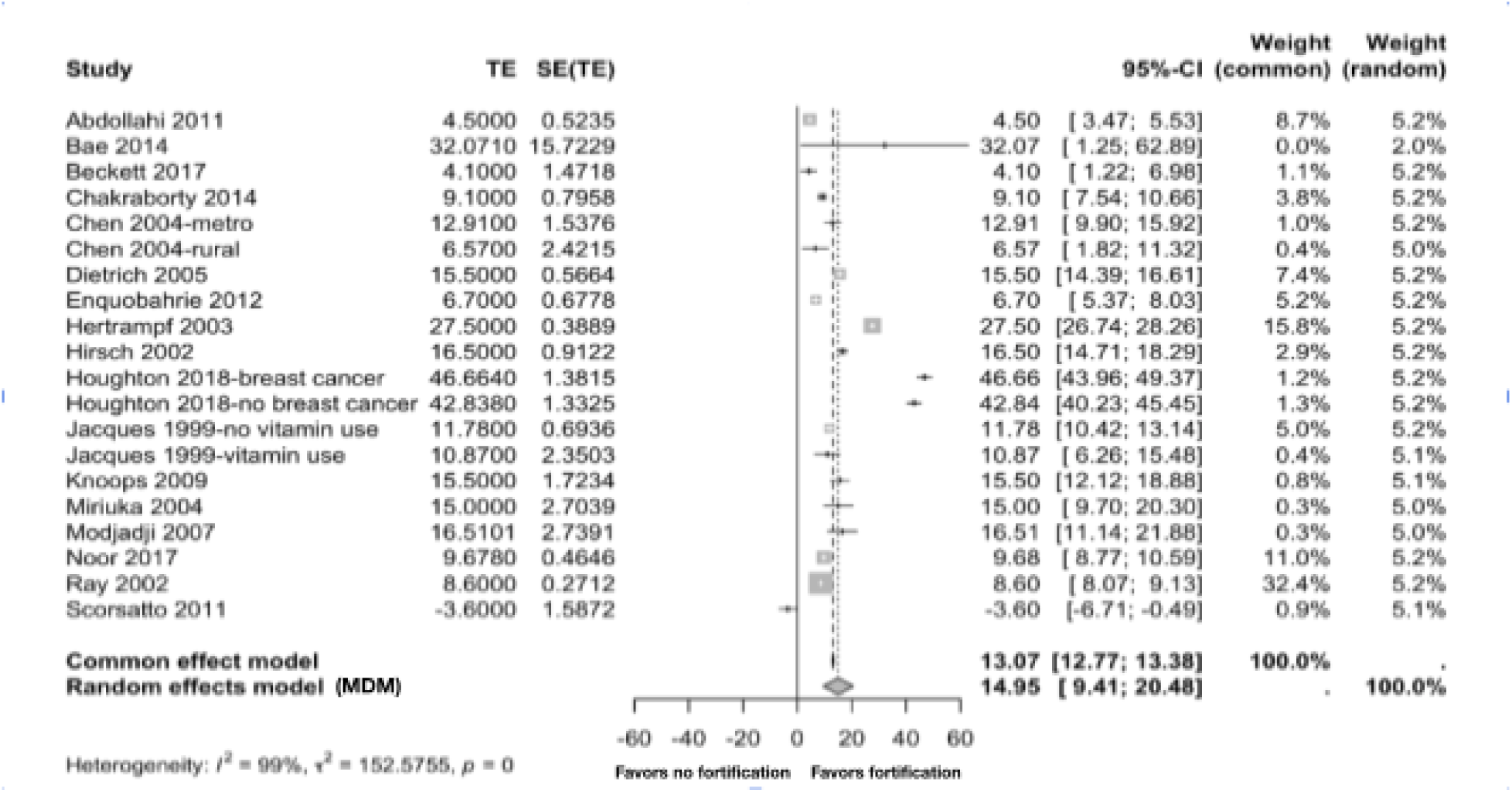
Changes in serum/plasma folate levels (nmol/L) following mandatory folic acid fortification of wheat flour, maize flour or rice in all population groups (n=17 studies). TE refers to the estimate of the treatment effect or standardized mean difference. SE refers to the standard error of the treatment effect or mean difference. MDM is the meta-differences of means from the random effects model.^1^ ^1^P-values less than or equal to zero are referred to as p<0.001 in the text.

These findings were supported by our analysis of prevalence outcomes, which found mandatory fortification of grains to be protective against both serum/plasma folate insufficiency and deficiency with RRs of 0.17 (95% CI 0.08-0.37) and 0.08 (95% CI 0.03-0.23), respectively (**Figure 3**, **Figure 4**). These results were similarly heterogenous for serum/plasma insufficiency (I^2^=100%, p<0.001) and deficiency (I^2^=99%, p<0.01). When prevalence of serum/plasma folate insufficiency was stratified by risk of bias (**Supplementary Figure 2**), studies with a high risk of bias had an mPR of 0.05 (95% CI 0.02-0.18), whereas studies with a low risk of bias had an mPR of 0.36 (95% CI 0.20-0.65). When prevalence of serum/plasma folate deficiency was stratified by risk of bias (**Supplementary Figure 3)**, studies with a high risk of bias had an mPR of 0.03 (95% CI 0.02-0.07), whereas studies with a low risk of bias had an mPR of 0.42 (95% CI 0.14-1.24).

**Figure 3:**
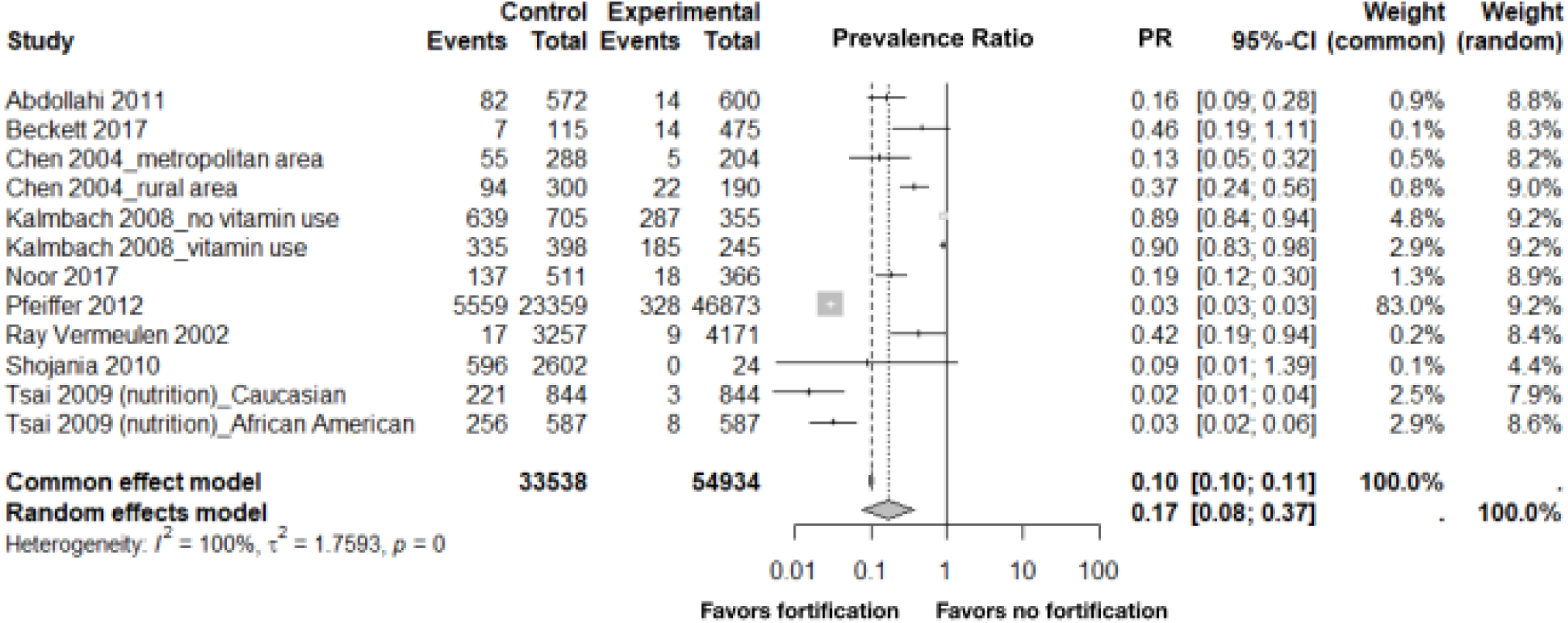
Changes in the prevalence of serum/plasma folate insufficiency following mandatory folic acid fortification of wheat flour, maize flour or rice in all population groups (n=9 studies). Event refers to a case of folate insufficiency, defined as <22.5 nmol/L, out of the total population at risk. Control refers to the individuals not exposed to mandatory folic acid fortification and experimental refers to individuals exposed to mandatory folic acid fortification.^1^ ^1^P-values less than or equal to zero are referred to as p<0.001 in the text.

**Figure 4:**
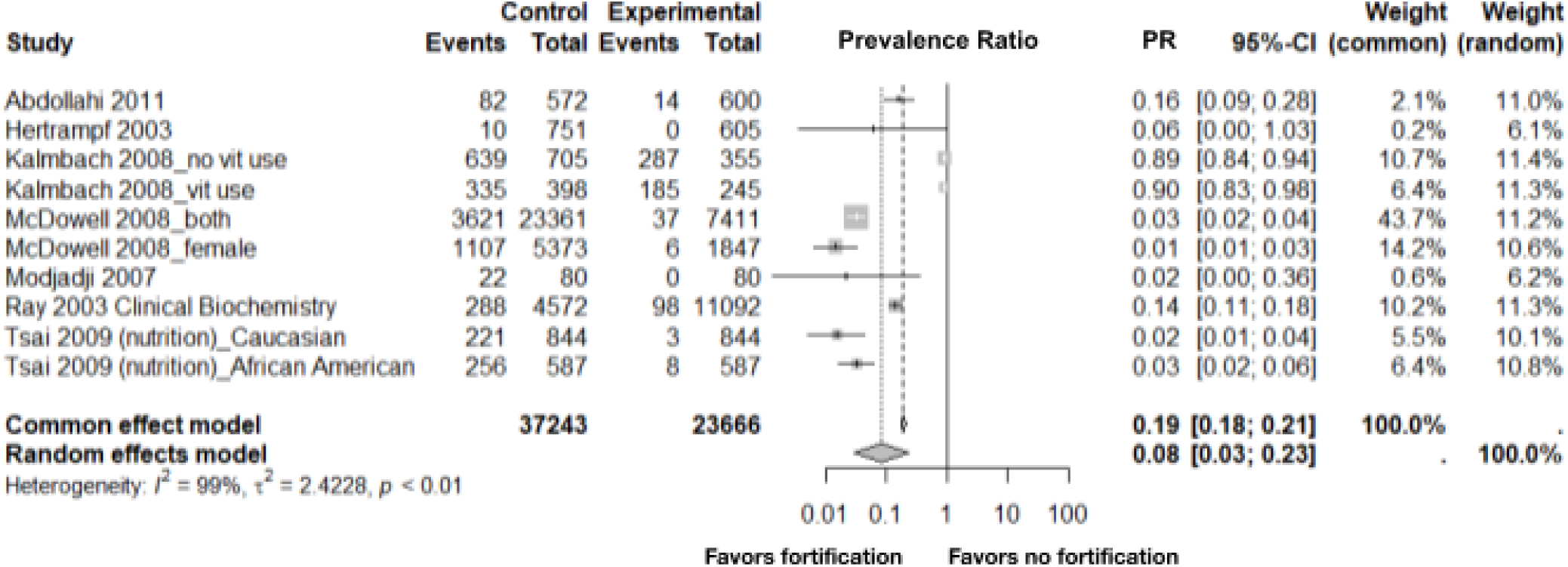
Changes in the prevalence of serum/plasma folate deficiency following mandatory folic acid fortification of wheat flour, maize flour or rice in all population groups (n=7 studies). Event refers to a case of folate deficiency, defined as <6.8 nmol/L, out of the total population at risk. Control refers to individuals not exposed to mandatory folic acid fortification and experimental refers to individuals exposed to mandatory folic acid fortification.^1^ ^1^P-values less than or equal to zero are referred to as p<0.001 in the text.

### Results of meta-analyses for RBC folate: continuous and binary outcomes

Mandatory fortification of wheat flour, maize flour, or rice with folic acid also led to an increase in RBC folate levels (**Figure 5**). The results of all studies included in the review were in the same direction although the magnitude of effect varied. In the overall analysis, the MDM was 329.4 nmol/L (95% CI 243.9-414.9), but the results were extremely heterogeneous with an I^2^ of 99%. When stratified by risk of bias (**Supplementary Figure 4**), studies with a high risk of bias had a MDM of 418.8 nmol/L (95% CI 223.8-613.8), whereas studies with a low risk of bias had a MDM of 289.7 nmol/L (95% CI 219.0-360.3).

**Figure 5:**
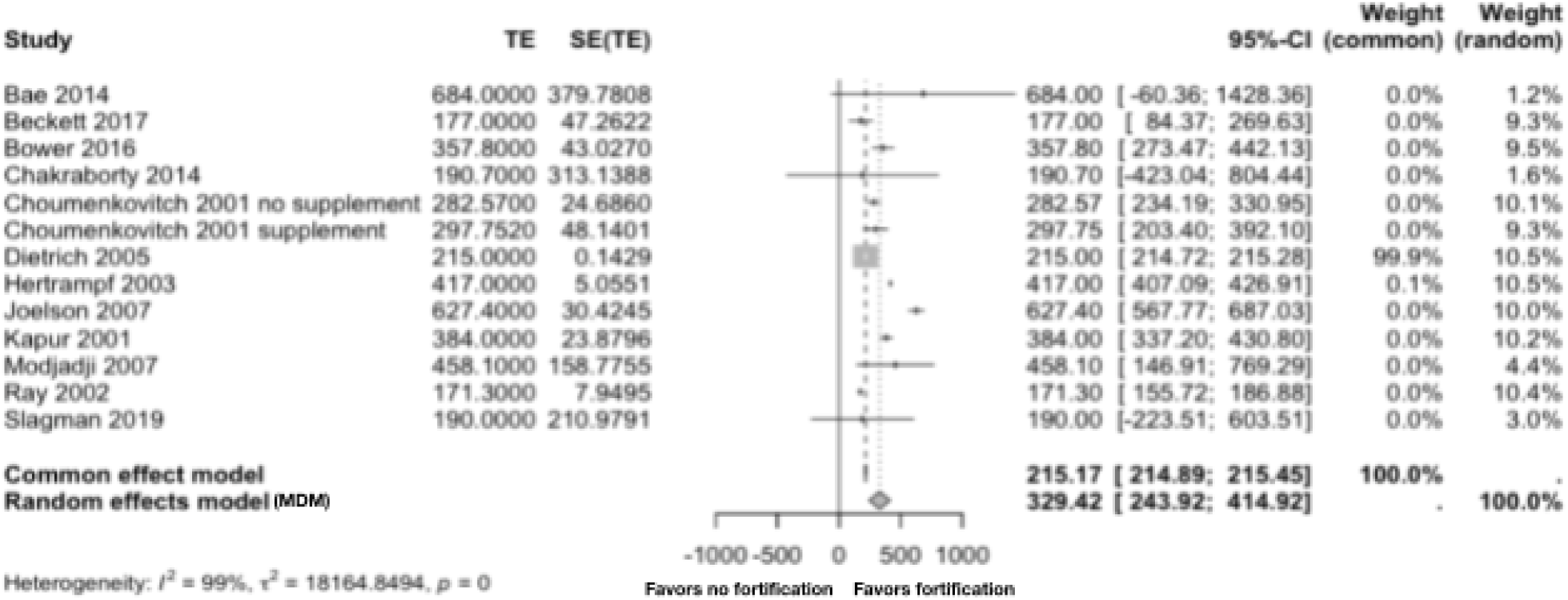
Changes in red blood cell (RBC) folate levels (nmol/L) following mandatory folic acid fortification of wheat flour, maize flour or rice in all population groups (n=12 studies). TE refers to the estimate of the treatment effect or standardized mean difference. SE refers to the standard error of the treatment effect or mean difference. MDM is the meta-differences of means from the random effects model.^1^ ^1^P-values less than or equal to zero are referred to as p<0.001 in the text.

These findings were aligned with our analysis of prevalence outcomes, which found mandatory fortification of grains to be protective against both RBC folate insufficiency and deficiency with mPR (95% CI) estimates of 0.16 (0.08-0.30) and 0.05 (0.01-0.46), respectively (**Figure 6**, **Figure 7**). These results for RBC folate insufficiency exhibited high heterogeneity (I^2^=97%, p<0.01), while those for RBC folate deficiency were less heterogeneous (I^2^=73%, p=0.03).

**Figure 6:**
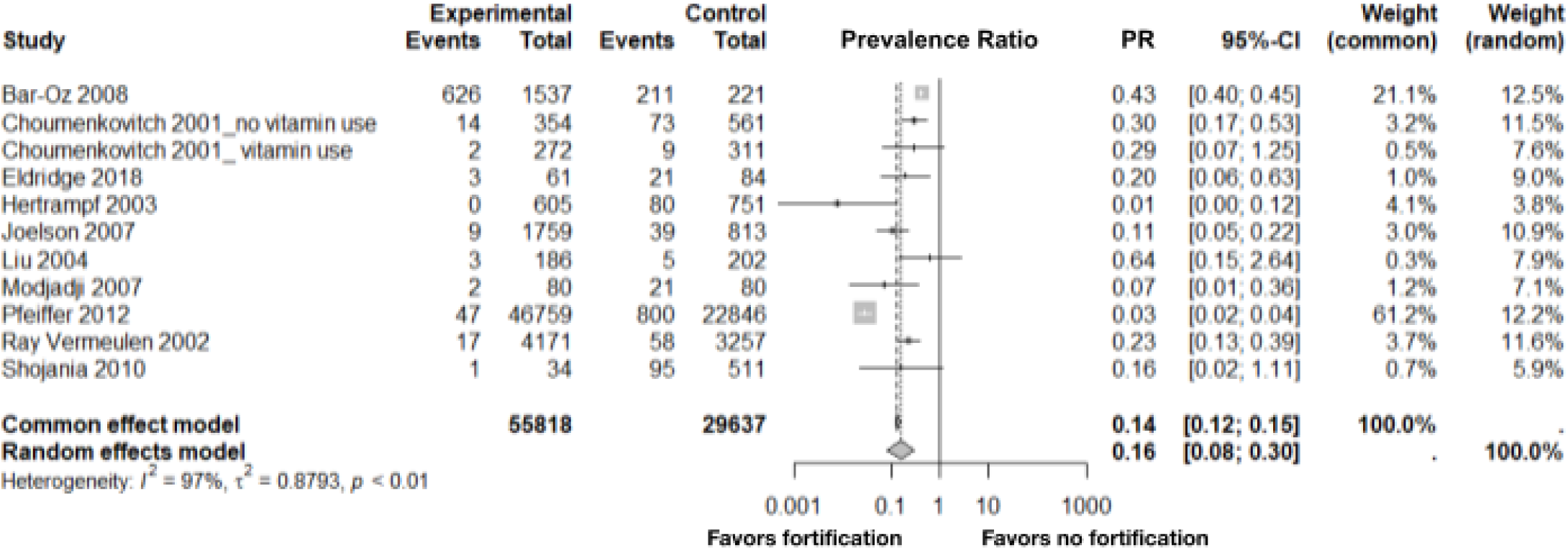
Changes in the prevalence of red blood cell (RBC) folate insufficiency following mandatory folic acid fortification of wheat flour, maize flour or rice in all population groups (n=10 studies). Event refers to a case of RBC folate insufficiency, defined as <906 nmol/L, out of the total population at risk. Control refers to the individuals not exposed to mandatory folic acid fortification and experimental refers to individuals exposed to mandatory folic acid fortification.^1^ ^1^P-values less than or equal to zero are referred to as p<0.001 in the text.

**Figure 7:**
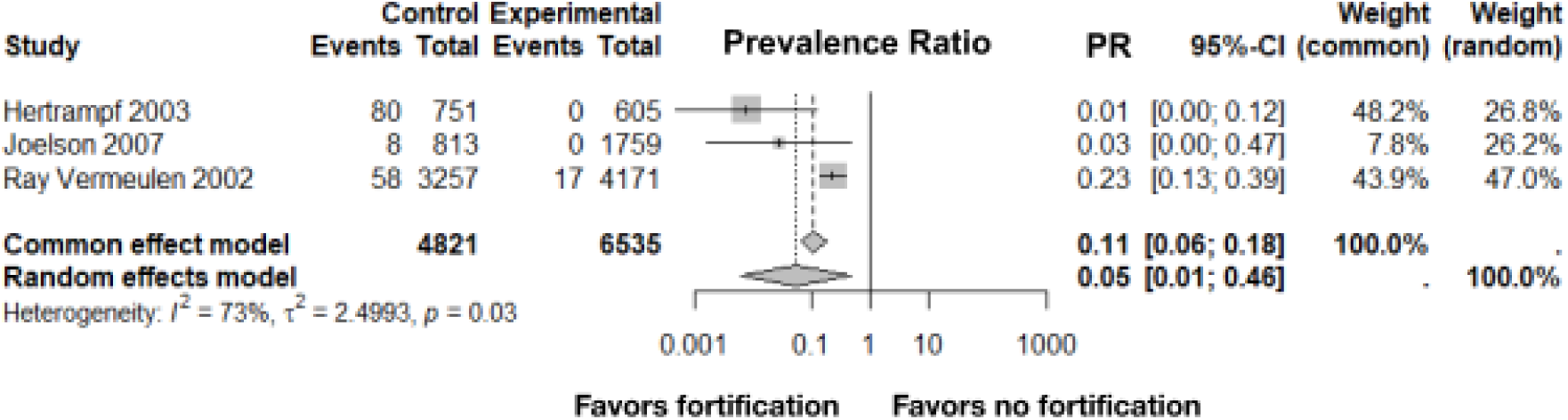
Changes in the prevalence of red blood cell (RBC) folate deficiency following mandatory folic acid fortification of wheat flour, maize flour or rice in all population groups (n=3 studies). Event refers to a case of RBC folate deficiency, defined as <226.5 nmol/L, out of the total population at risk. Control refers to individuals not exposed to mandatory folic acid fortification and experimental refers to individuals exposed to mandatory folic acid fortification.

When prevalence of RBC folate insufficiency was stratified by risk of bias (**Supplementary Figure 5)**, studies with a high risk of bias had an mPR of 0.13 (95% CI 0.06-0.28), whereas studies with a low risk of bias had an mPR of 0.33 (95% CI 0.21-0.53). The data were too sparse to stratify by risk of bias for prevalence of RBC folate deficiency.

### Biomarkers of excess folate status

Among the included studies, three reported biomarkers of excess folate status. Choumenkovitch et al. (2001)^46^ reported the prevalence of B-vitamin supplement users and non-users with red blood cell folate > 200 µg/L. Kalmbach and colleagues (2008)^47^ reported the prevalence of B-vitamin supplement users and non-users with serum/plasma folate >0.18 nmol/L and >1.35 nmol/L. Ray, Vermeulen, and collaborators (2002)^48^ reported the prevalence of adults with RBC folate >215 nmol/L and with serum/plasma folate >3.4 nmol/L.

### Publication bias

Our meta-analysis assessed the potential presence of publication bias using funnel plots and Egger’s test across both continuous and binary outcomes. For continuous outcomes, Egger’s test for serum/plasma folate level changes yielded a p-value of 0.60, indicating no significant publication bias (**Supplementary Figure 6**). Serum/plasma insufficiency also showed no significant bias (p = 0.33) (**Supplementary Figure 7**). In contrast, serum/plasma folate deficiency exhibited significant publication bias (p < 0.01), suggesting a higher likelihood of selective reporting in studies assessing this outcome (**Supplementary Figure 8**). RBC folate level changes showed no evidence of publication bias (p = 0.14) (**Supplementary Figure 9**). Similarly, Egger’s test for RBC insufficiency revealed no significant publication bias (p = 0.15) (**Supplementary Figure 10**). Finally, the number of studies (n = 3) reporting on RBC deficiency was too small to perform Egger’s test reliably, highlighting a limitation in sample size (**Supplementary Figure 11**).

## Discussion

To our knowledge, our study is the first systematic review and meta-analysis to evaluate the impact of mandatory grain fortification with folic acid on folate insufficiency, folate deficiency, and folate biomarker levels in populations worldwide. By synthesizing data from diverse settings, this study provides the most comprehensive evidence to date on the effectiveness of mandatory folic acid fortification policies. Specifically, the present meta-analysis comprehensively evaluates the impact of mandatory folic acid fortification of grains on population serum/plasma folate and RBC folate levels, as well as the prevalence of folate insufficiency and deficiency. While previous systematic reviews and meta-analyses have examined the effect of fortification on serum/plasma folate, none have assessed the impact on RBC folate.

Serum/plasma folate levels are highly sensitive to recent changes in dietary intake, making them a useful indicator of short-term folate status.^49^ RBC folate levels are not indicative of current dietary intake but rather reflect the longer-term folate status of the body.^49^ Together, serum/plasma folate and RBC folate results suggest that over both the short and the long term, grain fortification with folic acid improved population folate status. The reduced risk in serum/plasma folate and RBC folate insufficiency and deficiency serves as further evidence that this is an effective public health intervention. The results of almost all continuous studies (except for one (Scorsatto 2011^45^)) included in the review were in the same direction. Most studies showed no significant publication bias, except for serum/plasma folate deficiency, which demonstrated evidence of significant publication bias (p < 0.01). Overall, these findings align with previous research that has shown fortification to be effective in increasing folate levels, ultimately reducing the risk of neural tube defects.^3,9–14,30^ However, while the initial findings suggest a potential benefit of fortification, our assessment of publication bias suggests that findings related to serum/plasma folate deficiency may be influenced by selective reporting, warranting a more nuanced interpretation.

Previous systematic reviews and meta-analyses have primarily focused on the relationship between voluntary fortification or supplementation with folic acid and folate status outcomes.^14,27–29^ One 2013 systematic review and meta-analysis of three studies examined the effect of voluntary folate fortification on serum/plasma and RBC folate levels among women of reproductive age and reported increases in both outcomes.^28^ Both analyses had considerable heterogeneity^28^, but align with our findings that serum/plasma folate and RBC folate levels increase after fortification.

A 2019 systematic review and meta-analysis of 97 serum/plasma folate and 23 RBC folate studies – mostly intervention studies and supplementation trials – examined the dose-response relationship between folic acid and folate status levels.^27^ Similar to our results, they found that after initiation of folic acid intake, both serum/plasma folate and RBC folate concentrations increase.

A 2015 systematic review summarized data on 10 countries from the Latin American and the Caribbean region.^50^ Specifically, they examined pre- and post-fortification serum/plasma folate levels and found reductions in folate deficiency.^50^ While these results are from a more concentrated geographical region than ours, they corroborate our findings that fortification has successfully improved serum/plasma folate status globally.

Further, a more recent systematic review and meta-analysis of eight studies – RCTs, prospective cohorts, before-after, and cross sectional studies – focused on the impact of large-scale food fortification on serum/plasma folate levels among women of reproductive age primarily in LMICs.^14^ Most included studies examined mandatory fortification, but some were voluntary. The results of that meta-analysis were expressed as standardized mean differences and cannot be directly compared to our findings that are presented in terms of absolute differences; however both that study and our study reported a significant increase in folate levels (nmol/L) (e.g. SMD: 1.25 (95% CI: 0.50, 1.99) in Keats et al.) and both observed considerable heterogeneity across effect estimates (e.g. I^2^ : 99% in Keats et al.)^14^

Our findings are also consistent with the conclusions of a Cochrane review on folic acid fortification efficacy composed of randomized controlled trials (RCTs), quasi-RCTs, non-RCTs, and prospective observational studies with a control group which, although not meta-analyzed, reported increases in serum/plasma folate and RBC folate concentrations following fortification of wheat and/or maize flour with folic acid.^27,29^ Our study found a significant increase in both serum/plasma folate and RBC folate levels in adults, with a MDM of 15.0 nmol/L (95% CI: 9.4–20.5) for serum/plasma folate and 329.4 nmol/L (95% CI: 243.9–414.9) for RBC folate.

These results are in the same direction as those reported in the Cochrane review, which found a mean difference of 27.00 nmol/L (95% CI: 15.63–38.37) for plasma folate and a mean difference of 0.66 nmol/L (95% CI: 0.13–1.19) for RBC folate in adult women of reproductive age in Canada consuming folic acid fortified wheat flour bread rolls^29^. While the magnitude of the increase in RBC folate observed in our study was substantially higher, the direction of effect was consistent across both studies.^29,51^ Despite differences in study design, the consistency in direction and magnitude supports the effectiveness of mandatory folic acid fortification in improving folate status.

In terms of the literature on mandatory folic acid fortification effectiveness, there have been no meta-analyses on RBC folate levels after fortification. However, in 2010, Berry and colleagues conducted a systematic review of serum and RBC folate levels before and after mandatory fortification of wheat flour alone or in combination with maize flour in Canada, Chile, Costa Rica and the USA between 1988-2004.^52^ All studies and countries included in Berry’s review were also captured in our literature search, but not all met the final inclusion criteria for the meta-analysis. Similar to our study, Berry found an increase in both folate outcomes, regardless of country or study population.^52^

When stratified by risk of bias, studies with a low risk of bias had a higher increase in serum/plasma folate levels compared to those with a high risk of bias. However, studies with a low risk of bias had a lower increase in RBC folate levels compared to those with a high risk of bias. Prevalence ratios were greater for low risk of bias studies compared to high risk of bias studies when reporting on the prevalence of serum/plasma folate insufficiency and deficiency and the prevalence of RBC folate insufficiency. There was an insufficient number of studies with a low risk of bias reporting on RBC folate deficiency to conduct a meta-analysis stratified by bias risk. The findings suggest that the true effect of fortification with folic acid on folate status outcomes may be underestimated when all studies are included, although the 95% CIs overlapped between studies. This discrepancy highlights the importance of considering study quality and outcome measurement when interpreting the results.

Our meta-analysis assessed the potential presence of publication bias using funnel plots and Egger’s test across both continuous and binary outcomes. Publication bias was detected only for serum/plasma folate deficiency, while no significant bias was found for serum/plasma folate level changes, serum/plasma insufficiency, RBC folate level changes, or RBC insufficiency; assessment for RBC deficiency was inconclusive due to limited data. The presence of publication bias for serum/plasma folate deficiency suggests that the available evidence on this outcome may be skewed, potentially overestimating the prevalence or significance of deficiency due to selective reporting of positive or significant findings. This limits the reliability of conclusions drawn from this specific outcome and highlights the need for cautious interpretation, as well as further research using more comprehensive and transparently reported data. Two previous systematic reviews and meta analyses assessed publication bias.^29,53^ In both cases, this information was used to generate a risk of bias score.

Perhaps the most important limitation of the available literature is the pronounced heterogeneity of results across studies, due to varying population compositions and geographic regions. High heterogeneity makes it difficult to draw generalizable conclusions about the magnitude of effect from these findings.^44^ Additionally, the majority of studies were observational, limiting the ability to establish a direct, causal link to fortification efforts; such study designs are the norm when assessing the impact of mandatory governmental policies.^54^ This also raises the possibility of publication bias, a phenomenon in which studies with significant findings are more likely to be published,^55^ potentially overestimating the effect size. Evidence of possible publication bias was detected only in the analyses of serum/plasma folate deficiency, but not for serum/plasma folate level changes, serum/plasma insufficiency, RBC folate level changes, or RBC insufficiency (although assessment for RBC deficiency was inconclusive due to limited data).

Additionally, there is contrast in the techniques and their accuracy used to measure blood folate levels. As pointed out by Berry and colleagues, many studies used radioassay, which underestimates true blood folate by as much as 30%.^52^ Interpreting RBC folate results across studies can be challenging due to differences in measurement methods, as illustrated by Pfeiffer et al., who found that the microbiologic assay (MBA)—recommended by the CDC for its greater accuracy—yields higher RBC folate concentrations than older methods like the Bio-Rad radioassay, requiring adjustments to ensure comparability across time periods^56^. However, assuming techniques remained consistent between pre- and post-measurements, the differences in blood levels will carry the same bias. Researchers and decision-makers must take these limitations into consideration when using reviews to inform fortification policies, programs, and future research.

Despite these limitations, our study has several strengths, including the methodological rigor of a systematic review and statistical power of a meta-analysis. Our comprehensive literature search ensured all relevant studies were captured and standardized in regard to study type and quality, data extraction, and bias-risk assessment. The pooling of sample sizes and data from 31 different studies provides greater statistical power than individual studies can achieve on their own, producing more robust findings. Our analysis also allowed for the ability to stratify results by study quality (i.e., risk of bias) in an attempt to understand differences in effect. This research will strengthen the scientific evidence base on folic acid fortification and inform future research, policy and programs. Our review aims to provide an updated and comprehensive literature assessment on the association between mandatory fortification of grains (wheat flour, maize flour or rice) with folic acid and folate status in any country and population group.

In conclusion, this systematic review and meta-analysis offers the first quantitative assessment of the population-level impact of mandatory grain fortification with folic acid on both serum/plasma and RBC folate outcomes. By analyzing real-world data, we quantified the effectiveness of fortification in enhancing population folate status. Our findings demonstrate the significant public health benefits of these policies and programs in improving folate biomarkers and reducing folate insufficiency and deficiency globally. However, the observed heterogeneity and influence of study quality underscore the need for well-designed and -implemented fortification programs to maximize health outcomes.

## Data Availability

All data produced in the present work are contained in the manuscript

https://osf.io/r2gk4

## Acknowledgements

We would like to acknowledge Emory University librarian Sharon Leslie for her assistance in formatting the second search completed in PubMed and Embase. Her guidance helped us to ensure all possible studies were captured.

## Author contributions

HP conceived and designed the research, developed the overall plan, and oversaw the study. AK, JP, LC, and MEG conducted the experiments and collected data, while MG also provided essential materials. Data analysis and statistical evaluation were performed by JP and SH. The manuscript was primarily written by HP, JP, AK, LC, and MEG, with JP holding primary responsibility for the final content. All authors have read and approved the final manuscript.

## Disclaimer

Several co-authors are employed with Emory University and the Food Fortification Initiative. These organizations help country leaders promote, plan, implement, monitor or evaluate food fortification.

HIC: high-income country
LMIC: low or middle-income country
MDM: meta-difference of means
NTD: neural tube defect
OSF: Open Science Framework
PICO: population, intervention, comparison, outcome
RBC: red blood cell
ROB: risk of bias
PR: prevalence ratio

