## Supplementary Materials for "Impact of Mandatory Grain Fortification with Folic Acid on Population Folate Levels and the Risk of Folate Deficiency and Insufficiency: A Systematic Review and Meta-Analysis"

**Supplementary Material**

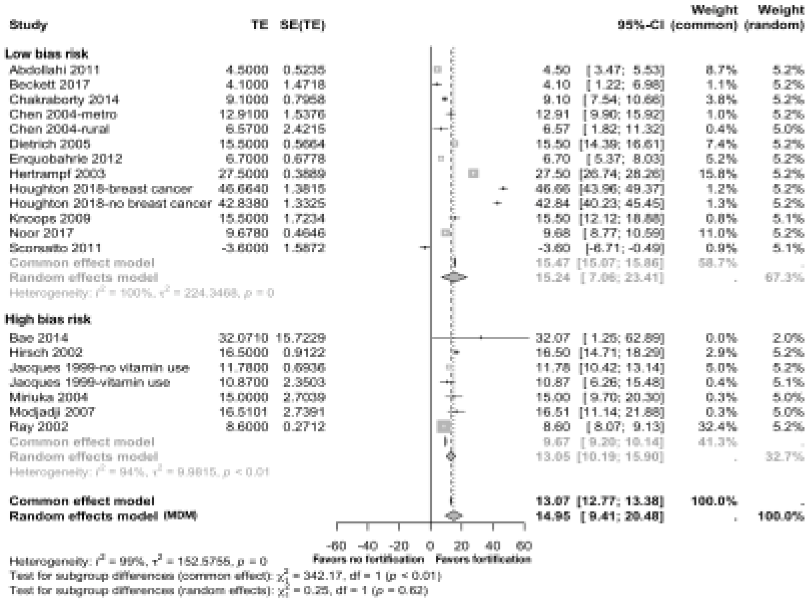


*Supplementary Figure 1*: Changes in serum/plasma folate levels (nmol/L) following mandatory folic acid fortification of wheat flour, maize flour or rice stratified by bias risk (n=17 studies). Bias risk categorization is based on the number of risk of bias assessment criteria met (**Table 3, Table 4**). Low bias risk: <3 criteria met; High bias risk: ≥3 criteria met. TE refers to the estimate of the treatment effect or standardized mean difference. SE refers to the standard error of the treatment effect or mean difference. MDM is the meta-differences of means from the random effects model.[1]

[1]P-values less than or equal to zero are referred to as p<0.001 in the text.

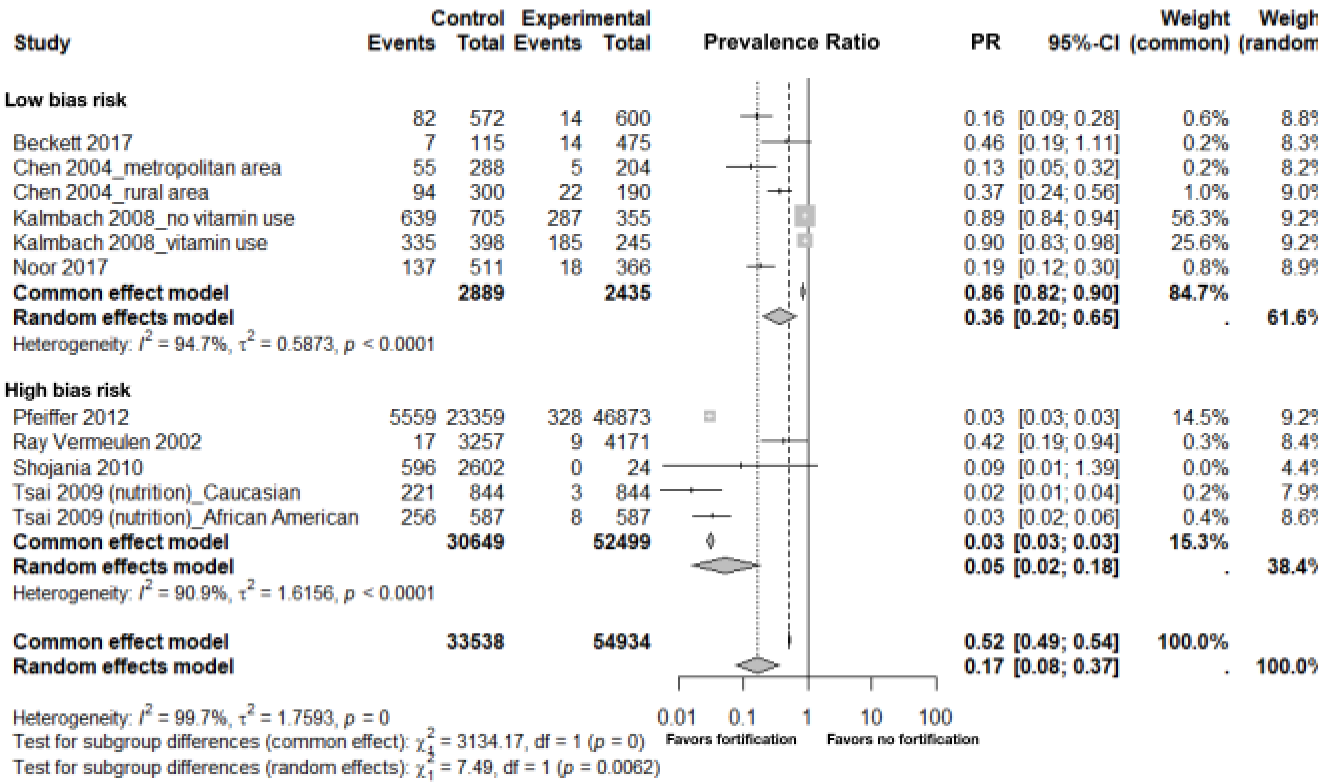


*Supplementary Figure 2*: Changes in the prevalence of serum/plasma folate insufficiency following mandatory folic acid fortification of wheat flour, maize flour or rice stratified by bias risk (n=9 studies). Bias risk categorization is based on the number of risk of bias assessment criteria met (**Table 3, Table 4**). Low bias risk: <3 criteria met; High bias risk: ≥3 criteria met. Event refers to a case of folate insufficiency, defined as <22.5 nmol/L, out of the total population at risk. Control refers to the individuals not exposed to mandatory folic acid fortification and experimental refers to individuals exposed to mandatory folic acid fortification.[1]

[1]P-values less than or equal to zero are referred to as p<0.001 in the text.

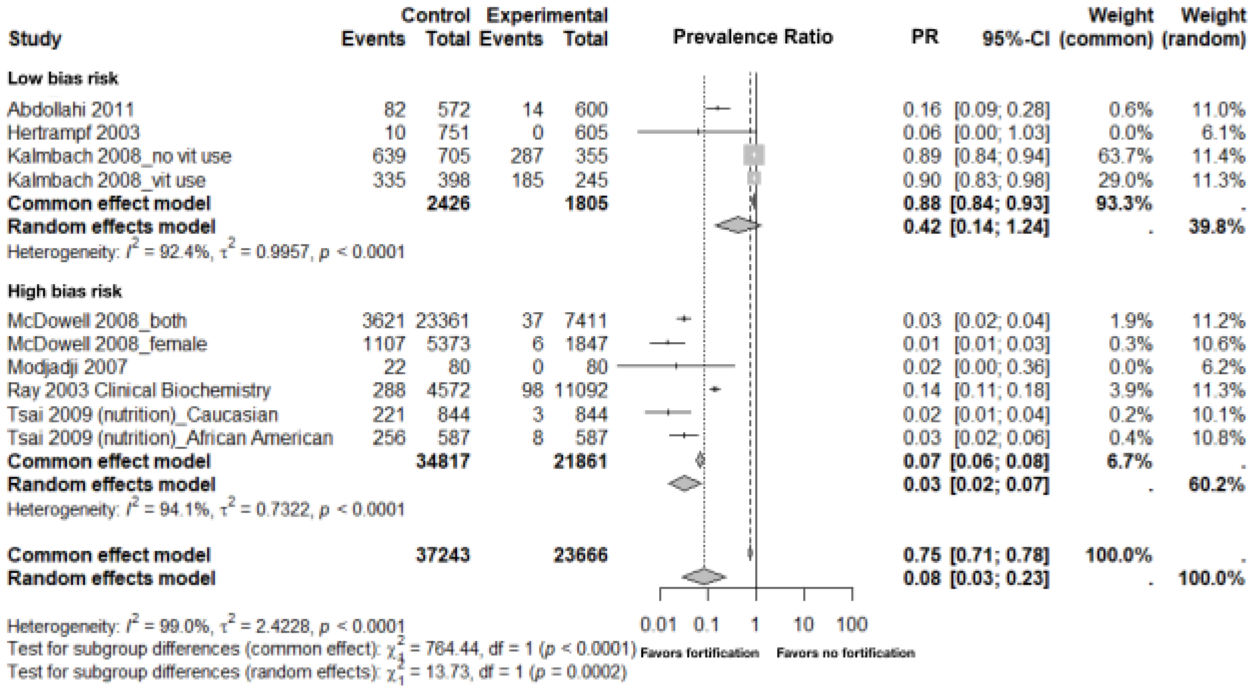


*Supplementary Figure 3*: Changes in the prevalence of serum/plasma folate deficiency following mandatory folic acid fortification of wheat flour, maize flour or rice stratified by bias risk (n=7 studies). Bias risk categorization is based on the number of risk of bias assessment criteria met (**Table 3, Table 4**). Low bias risk: <3 criteria met; High bias risk: ≥3 criteria met. Event refers to a case of folate deficiency, defined as <6.8 nmol/L, out of the total population at risk. Control refers to individuals not exposed to mandatory folic acid fortification and experimental refers to individuals exposed to mandatory folic acid fortification.[1]

[1]P-values less than or equal to zero are referred to as p<0.001 in the text.

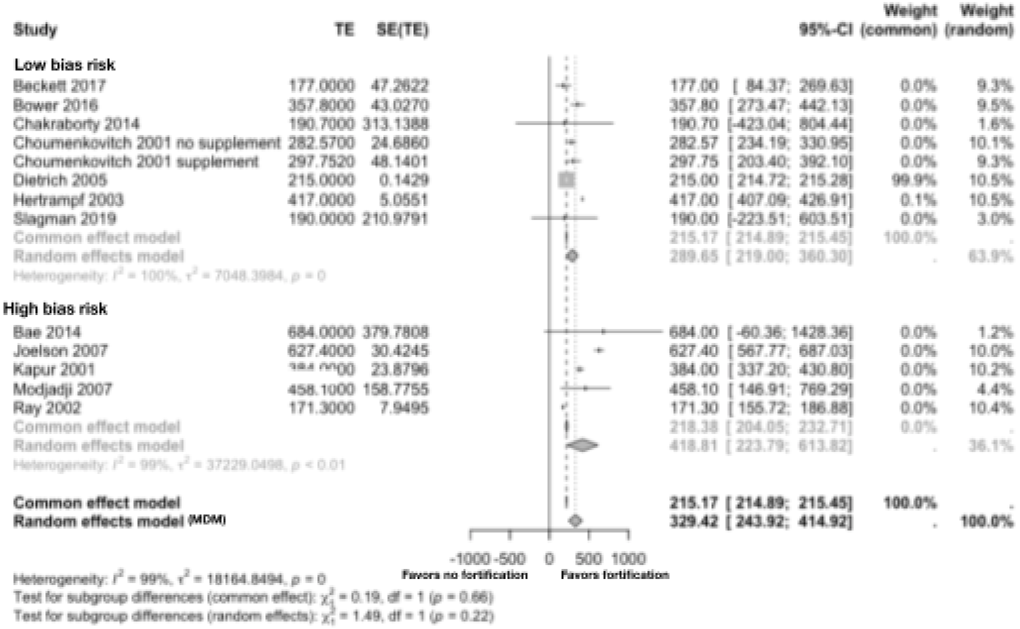


*Supplementary Figure 4*: Changes in red blood cell (RBC) folate levels (nmol/L) following mandatory folic acid fortification of wheat flour, maize flour or rice stratified by bias risk (n=12 studies). Bias risk categorization is based on the number of risk of bias assessment criteria met (**Table 3, Table 4**). Low bias risk: <3 criteria met; High bias risk: ≥3 criteria met. TE refers to the estimate of the treatment effect or standardized mean difference. SE refers to the standard error of the treatment effect or mean difference. MDM is the meta-differences of means from the random effects model.[1]

[1]P-values less than or equal to zero are referred to as p<0.001 in the text.

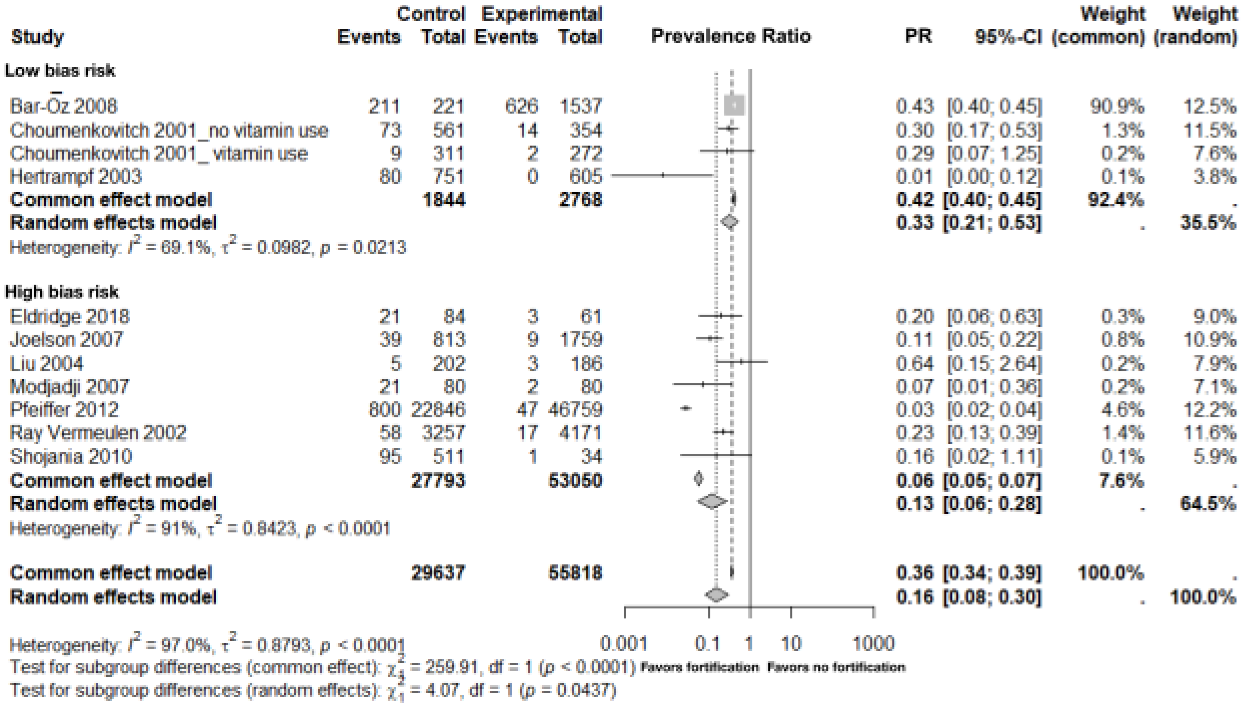


*Supplementary Figure 5:* Changes in the prevalence of red blood cell (RBC) folate insufficiency following mandatory folic acid fortification of wheat flour, maize flour or rice stratified by bias risk (n=10 studies). Bias risk categorization is based on the number of risk of bias assessment criteria met (**Table 3, Table 4**). Low bias risk: <3 criteria met; High bias risk: ≥3 criteria met. Event refers to a case of RBC folate deficiency, defined as <226.5 nmol/L, out of the total population at risk. Control refers to individuals not exposed to mandatory folic acid fortification and experimental refers to individuals exposed to mandatory folic acid fortification.[1]

[1]P-values less than or equal to zero are referred to as p<0.001 in the text.

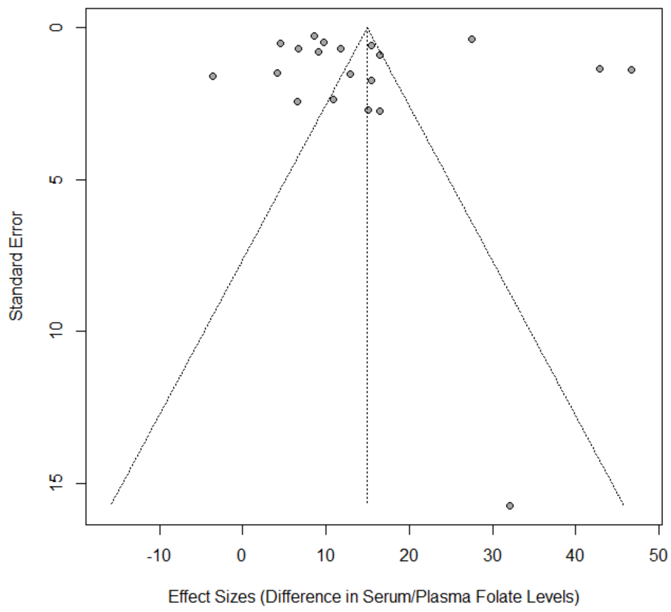


*Supplementary Figure 6*: Funnel plot of serum/plasma folate level changes (Egger's test p-value = 0.60).

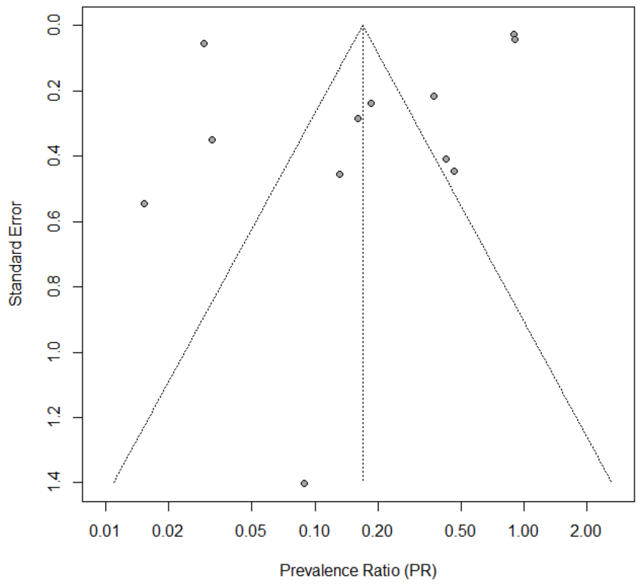


*Supplementary Figure 7*: Funnel plot of serum/plasma folate insufficiency pre vs. post fortification (Egger's test p-value = 0.33).

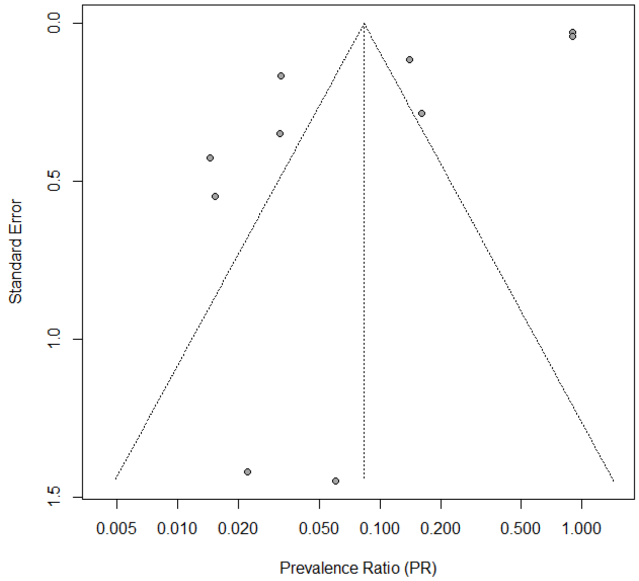


*Supplementary Figure 8*: Funnel plot of serum/plasma folate deficiency pre vs. post fortification (Egger's test p-value < 0.01).

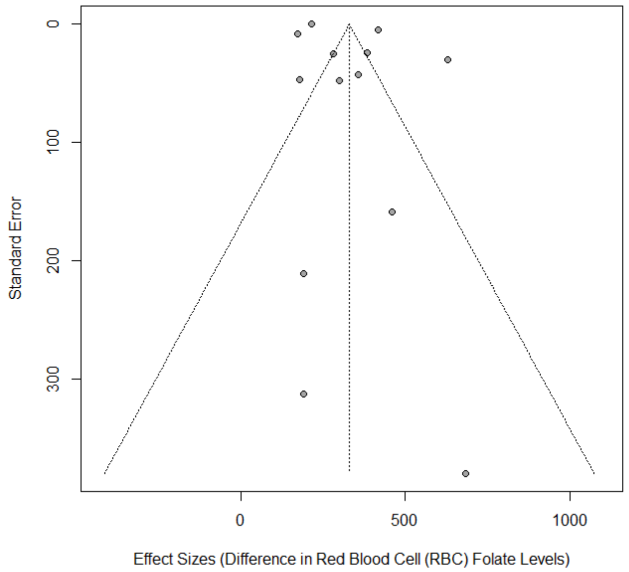

*Supplementary Figure 9:* Funnel plot of red blood cell (RBC) folate level changes (Egger's test p-value = 0.14).

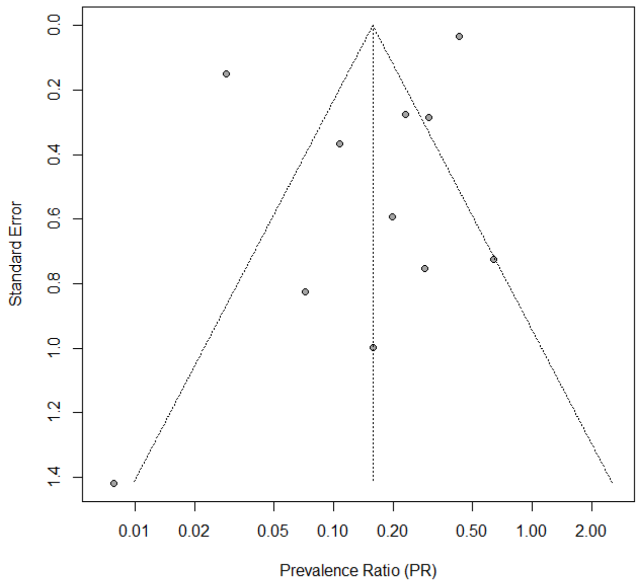


*Supplementary Figure 10*: Funnel plot of red blood cell (RBC) insufficiency pre vs. post fortification (Egger's test p-value = 0.15).

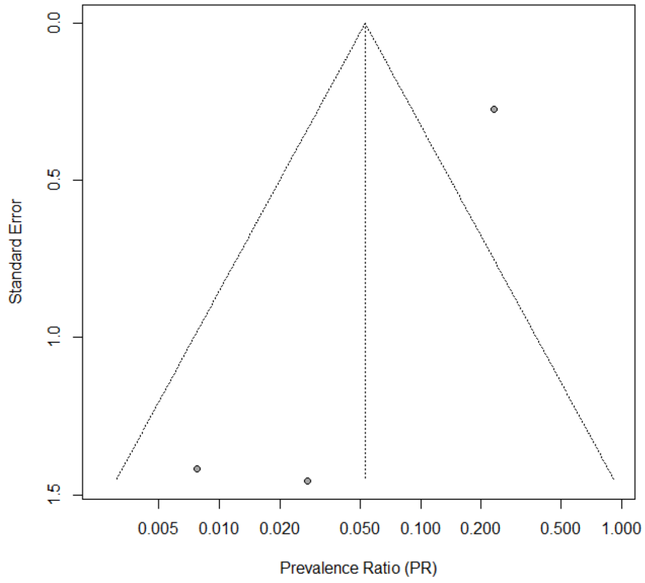


*Supplementary Figure 11*: Funnel plot of red blood cell (RBC) deficiency pre vs. post fortification. No Egger's test was conducted for RBC deficiency due to the small sample size (n = 3 studies).
